# Aggregated genomic data as cohort-specific allelic frequencies can boost variants and genes prioritization in non-solved cases of inherited retinal dystrophies

**DOI:** 10.1101/2022.05.09.22274851

**Authors:** Ionut-Florin Iancu, Irene Perea-Romero, Gonzalo Núñez-Moreno, Lorena de la Fuente, Raquel Romero, Almudena Ávila-Fernandez, María José Trujillo-Tiebas, Rosa Riveiro-Álvarez, Berta Almoguera, Inmaculada Martín-Mérida, Marta Del Pozo-Valero, Alejandra Damián-Verde, Marta Cortón, Carmen Ayuso, Pablo Minguez

## Abstract

The introduction of NGS in genetic diagnosis has increased the repertoire of variants and genes involved and the amount of genomic information produced. We built an allelic-frequency (AF) database for a heterogeneous cohort of genetic diseases to explore the aggregated genomic information and boost diagnosis in inherited retinal dystrophies (IRD). We retrospectively selected 5683 index-cases with clinical exome sequencing tests available, 1766 with IRD and the rest, with diverse genetic diseases. We calculated subcohort’s IRD specific AF and compare it with suitable pseudocontrols. For non-solved IRD cases, we prioritized variants with a significant increment of frequencies, with 8 variants that may contribute to explain the phenotype, and 10/11 of uncertain significance that were reclassified as likely-pathogenic according to ACMG. Besides, we developed a method to highlight genes with more frequent pathogenic variants in IRD cases than in pseudocontrols weighted by the increment of benign variants in the same comparison. We identified 18 genes for further studies that provided new insights in five cases. This resource can also help to calculate the carrier-frequency in IRD genes. A cohort-specific AF database assist with variants and genes prioritization and operate as an engine that provides new hypothesis in non-solved cases, augmenting diagnosis rate.

## 1. Introduction

Rare diseases are chronically debilitating or life-threatening, and have a prevalence in Europe of less than 1 of every 2000 people [1]. Inherited retinal dystrophies (IRD) are a group of rare diseases with a degenerative and progressive course and are caused by primary affection of photoreceptors and retinal pigmentary epithelial [2]. All together they affect 1 of every 3000-4000 people in the western world [3]. They are clinically heterogeneous, covering several syndromes (e.g., Usher, Bardet-Biedl -BBS-, or Joubert) [4–6], as well as non-syndromic forms as retinitis pigmentosa [2] and macular dystrophies [7]. They have overlapped phenotypes and display any form of inherited patterns.

During the last two decades, next generation sequencing (NGS) techniques have transformed research on genetic rare diseases with a substantial increase in volume of available genomic data and knowledge generated [8,9]. Although several sequencing tests are available, a widespread approach in genetic diagnosis is to sequence the coding region of known clinically relevant genes (∼4500), the so-called clinical exome (CE). A CE test detects several thousand variants, which need to be filtered and prioritized in order to highlight those responsible of the phenotype [10]. Regarding the task of filtering, in low prevalent diseases, apart from quality filters, a low population frequency is one of the first requirements to purge non-causal variants [11]. Several global genomic initiatives [12,13] provide allele frequencies on large populations, although frequencies from local cohorts provide a better estimation of real variant prevalence [14,15] and have proven to identify rare pathogenic variants [15,16].

In the absence of genomic information of a priori healthy people, Mendelian diseases can provide a good estimate of allele frequencies in the general population as pseudocontrols (PC) for other non-related diseases [17,18]. In the same terms, PC can also be applied for the calculation of carrier frequencies (CF) [16,19] of causal variants of non-related diseases in genes with a recessive inheritance pattern, as well as in the analysis of trios [18,19]. In more complex scenarios, where modifying and risk/protective variants may tune the effect of causal variants, the mutational landscape of a disease may help identifying: i) genetic pleiotropy together with causal variants in recessive forms [22], ii) digenic inheritance [23], or iii) disease-associated triallelic sites as in BBS [24].

With all these premises, we hypothesize that a database of variant allele frequencies calculated over a heterogeneous cohort of genetic diseases enriched in IRD cases can help to improve the detection of previously unnoticed, underrated or unknown causal variants and gene-disease associations. Additionally, it can assist to uncover disease cases with overlapping phenotypes, as well as to describe the carrier frequencies of recessive variants. Thus, we built a database with the genomic data of a large cohort with various genetic diseases and developed methods to compare IRD-specific and PC frequencies. This tool is used as a global reanalysis platform to study frequent variants and over-mutated genes in IRD non-solved cases.

## 2. Results

### 2.1 A multi-disease cohort database of variant frequencies to study the aggregated signal in IRD genomic landscape

We compiled a heterogeneous cohort of 5683 patients with genetic diseases referred to the Genetics Department of the UH-FJD and with a clinical exome sequencing test available (see Methods). The cases were distributed into three groups of diseases as: inherited retinal dystrophies (IRDs), with 1766 cases, other eye-related diseases (OERDs) with 386 cases, and non-eye related diseases (NRD), 3531 cases (Figure 1A). Additionally, IRD cases were classified according to their diagnostic status as: solved (n=955 cases, 54%) and non-solved (n=811 cases, 46%). Within the cohort of non-solved IRD patients, we had 447 cases with no candidate variants (25% of the total), that is, excluding cases with a pathogenic/likely pathogenic variant reported in recessive cases (named partially solved cases), and cases with 1-2 VUS reported in dominant/recessive cases respectively (named VUS cases), (Figure 1B).

**Figure 1.**
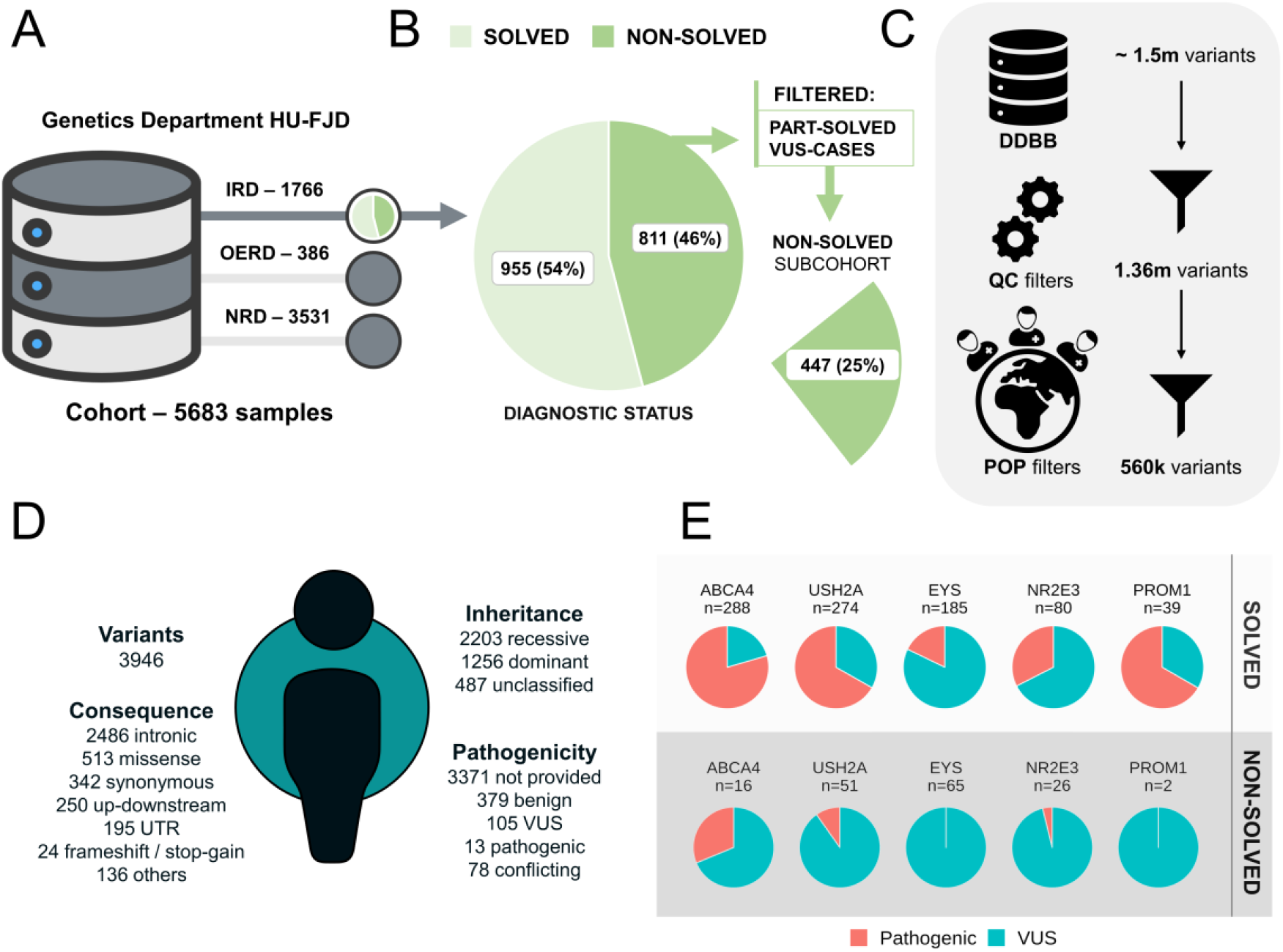
A heterogeneous cohort of rare diseases with the diagnostic status and variant composition in IRD cases. A) The cohort of patients with suspected rare genetic diseases at the Genetics Department of HU-FJD was divided into three subcohorts. An IRD subcohort of 1766 samples, an Other Eye Related Diseases (OERD) of 386 cases and a pseudocontrol subcohort of non-eye related diseases (NRD) with 3531 samples. B) IRD diagnostic status of the samples included in IRD subcohort was: solved and non-solved. The subcohort of non-solved IRD cases with no candidate variants represent the 25%. C) Flow chart of different filters applied to variants according to quality control (QC) and population (POP) filters. D) Summary of the variants included in the database in an average IRD case, values represent the average of all IRD samples. E) Proportion of pathogenic and VUS variants detected in IRD cases in the genes with more pathogenic variants in IRD solved cases, top 5 genes are shown.

CEs were reanalyzed and variants detected and extracted for each case. We performed a quality filter and removed variants with a potential sub-population bias between IRD and other cases (Figure 1C and Methods). Thus, for the 5683 samples together, around 560K unique variants in 5046 genes were left for further analyses.

Using this set of variants, an average IRD individual have approximately 4k non-polymorphic variants (Allele frequency, AF<0.1) within the specified sequenced region (Figure 1D), being mostly intronic (N=2486, 63%), missense (N=513, 13%), and synonymous (N=342, 9%). According to the inheritance pattern observed for the genes based on their associated diseases, 2203 variants (56%) have a recessive pattern, 1256 (32%) dominant, and 487 (12%) either without a clear pattern, undetermined, or X-linked. Regarding their pathogenicity, 379 (10%) are benign or likely benign, 105 (3%) VUS, and 13 (0,3%) pathogenic or likely pathogenic (according to ClinVar). However, there are still a large percentage of variants with missing or conflicting annotation (N=3449, 87%). From a cohort perspective, the number of pathogenic (including likely pathogenic) variants and VUS are unequally distributed over IRD associated genes when comparing solved-IRD and non-solved-IRD cases (Figure 1D). Top 5 genes with more pathogenic variants (including likely pathogenic) in solved-cases have a greater ratio pathogenic variants / VUS for IRD solved cases than for IRD non-solved cases.

### 2.2 IRD-specific highly frequent variants

In the diagnosis of rare diseases using NGS data, variants must be prioritized in order to facilitate the detection of the causal mutations within the large amount of variation found in a single experiment. Here we aimed to test if the comparison between the frequency of variants in cases and controls was able to extract deleterious variants. Thus, we focused in the IRD subcohort and treated solved and non-solved cases separately. Partially solved and VUS cases were excluded from non-solved subcohort in order to work with a subcohort with no candidate variants. As controls for both subcohorts we used the set of patients with non-related diseases, from now on called pseudocontrols (PC) (See Methods). Allele frequencies (AF), allele numbers (AN) and allele counts (AC) were calculated for solved-IRD cases, non-solved-IRD cases (free of candidate variants) and PC cases.

Next, for both, solved and non-solved IRD cases independently, we compared in every variant, its AF (solved-AF or non-solved-AF) with its AF in the PC subcohort (PC-AF), see Methods. We defined the “most frequent variants” in a IRD subcohort (IRD-MFVs) as those within the top 10% with the highest log2 of the fold change, log2(FC) values in the comparison performed. The cut offs for the log2(FC) were 3.12 and 3.92 in solved and non-solved IRD cases respectively (Figure 2A-B). The distribution of log2(FC) values in solved and non-solved IRD cases is shown in the Supplementary Figure S1. MFVs are prioritized for a posterior reevaluation of cases. Non-prioritized variants are defined as those with a higher frequency in IRD (FC>0) but below the significant threshold. Classifying IRD-MFVs according to their clinical relevance and removing those not informative (see Methods), we found IRD-MFVs enriched in deleterious variants in both solved and non-solved cases compared to non-prioritized variants (Fisher’s exact test, p-values=4.77E-56 and 1.69E-32, respectively; Figure 2C-D and Supplementary Tables S1-2).

**Figure 2.**
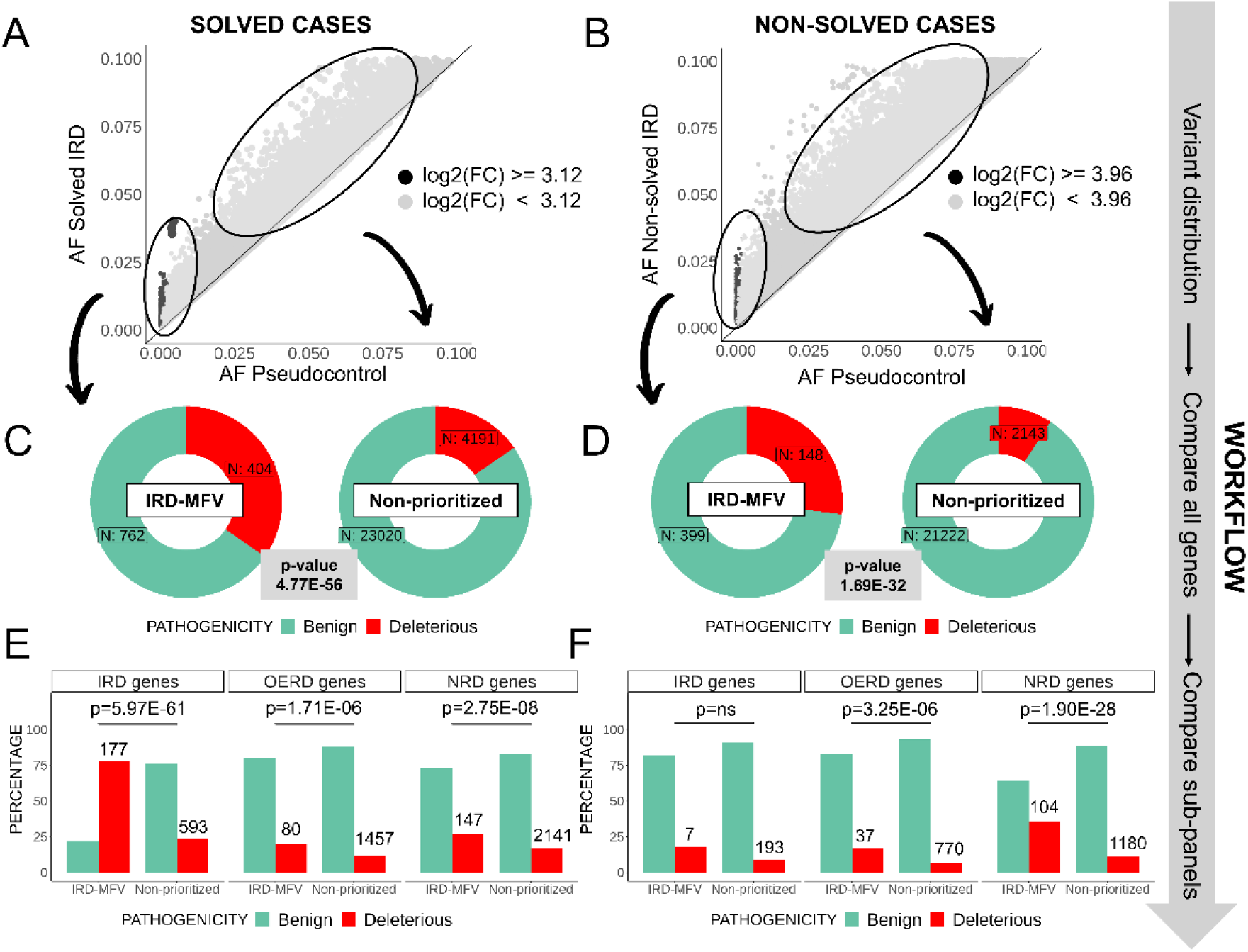
Comparison of the percentage of deleterious variants in prioritized and non-prioritized variants in solved and non-solved IRD patients. Variant AFs are compared in inherited retinal dystrophies (IRD) solved (A) and non-solved (B) subcohorts against pseudocontrols (PC) using fold changes (FC). IRD more frequent variants (IRD-MFVs), highlighted in dark, were defined using FC thresholds at .90 percentiles of all FC in each comparison (A-B). Proportion of deleterious and benign variants in both solved (C) and non-solved IRD cases (D) and the p-values representing the enrichment of deleterious variants in IRD-MFVs. Enrichment analyses are also performed dividing the IRD-MFVs according to the genes in which they are located, grouping them in: IRD genes, other eye related diseases (OERD genes) and other non-eye related diseases (NRD genes) (E-F). Total number of deleterious variants in each group is noted at the top of the red bars. Non-significant p-values are marked as “ns”.

Focusing on the type of genes where the IRD-MFVs are located, we divided IRD-MFVs as present in IRD-associated genes, OERD-related genes, and NRD-associated genes. In IRD-solved cases, regarding IRD-MFVs in IRD-genes only, the ratio deleterious / benign is 78% (177/256), that is significantly higher than the same proportion in non-prioritized variants (14%, 593/1926, Figure 2E, Supplementary Table S1). A different trend was observed in non-solved IRD cases where we found no significant differences in the percentages of deleterious/benign variants in IRD-genes between IRD-MFVs and non-prioritized variants (Figure 2F, Supplementary Table S2). In IRD-MFVs in OERD-genes, we found also more deleterious variants in our prioritized set in both, solved and non-solved cases (Fisher’s exact test, p-value=1.71E-06 and p-value=3.25E-06, respectively; Figure 2E-F, Supplementary Tables S1-2). Finally, we also observed an enrichment of deleterious variants in the IRD-MFVs located in NRD-genes in solved and non-solved IRD cases (p-value=2.75E-08 and p-value=1.90E-28, respectively; Figure 2E-F, Supplementary Tables S1-2).

Furthermore, solved and non-solved cases were divided into disease sub-categories as syndromic, non-syndromic and macular dystrophy forms (Supplementary Figure S2), and the analysis was repeated for each sub-group. Thus, as for all solved cases considered as a whole; syndromic, macular dystrophies and non-syndromic forms behave very similarly, with more deleterious variants in the prioritized sets for IRD genes (Supplementary Figure S3A-C, Supplementary Tables S3-4). For non-solved cases, we observed an increase in deleterious variants in the prioritized sets for macular dystrophy and non-syndromic forms (Supplementary Figure S3D-F and Supplementary Tables S3-4).

Next, we performed a reevaluation of non-solved IRD cases carrying an IRD-MFV and found eight variants that provide additional insights in 8 cases (Table 1). In this reevaluation, the diagnosis status of the cases is classified as: 1) “solved”, when the case has a conclusive diagnosis thanks to the variant(s) identified, or 2) “non-solved”, in any other case. Non-solved cases were annotated according to findings that may help in a future diagnosis as: 2.1) “partially solved”, if a heterozygous pathogenic or likely pathogenic variant is found within a recessive gene that fits the observed phenotype. And 2.2) “with evidence”, when the variants identified in the analysis are: 2.2.1) pathogenic/likely pathogenic variants in genes not yet associated with the disease but with some evidence published in the literature or with overlapping phenotypes, or 2.2.2) VUS in a gene associated to the phenotype when one VUS in dominant or 1-2 VUS in recessive genes were found.

**Table 1.** Identification of most frequent variants in IRD (IRD-MFV) allowed the reevaluation of non-solved IRD cases. A total of 8 cases (one per row) gained knowledge in their reassessment. The gene panel column refers to the type of diseases that the gene has been associated with, inherited retinal dystrophies (IRD), other eye related diseases (OERD) and other non-eye related diseases (NRD). In the phenotype, abbreviations are RCD: Rod-cone dystrophy, RP: retinitis pigmentosa and MD: macular dystrophy. In variant column: HGVSc, HGVSp and ACMG classification are included. In “Other Variants”, for those biallelic cases, the second variant found after reviewing the case is included. Genotype column has information about the genotype of the variant(s) in the sample. Diagnostic status column has the new diagnostic status of the case after the reanalysis, as: 1) “solved”, when the case has a conclusive diagnosis thanks to the variant(s) identified, or 2) “non-solved”, in any other case. Non-solved cases were annotated according to findings that may help in a future diagnosis as: 2.1) “partially solved”, if a heterozygous pathogenic or likely pathogenic variant is found within a recessive gene that fits the observed phenotype. And 2.2) “with evidence”, when the variants identified in the analysis are: 2.2.1) pathogenic/likely pathogenic variants in genes not yet associated with the disease but with some evidence published in the literature or with overlapping phenotypes, or 2.2.2) VUS in a gene associated to the phenotype when one VUS in dominant or 1-2 VUS in recessive genes were found.

| Gene | Gene panel | Phenotype | Prioritized Variant (HGVS, HGVS, ACMG classification) | Other variants | Genotype | Diagnostic status |
| --- | --- | --- | --- | --- | --- | --- |
| *CDH23* | IRD | RCD | NM_022124.6: c.6050-9G>A<br>pathogenic | - | Monoallelic 0/1 | Partially solved |
| *MYO7A* | IRD | Usher syndrome | NM_000260.4: c.1996C>T<br>(p. Arg666Ter);<br>pathogenic | NM_000260.4: c.3764del<br>(p. Lys1255ArgfsTer8)<br>(pathogenic) | Biallelic 0/1, 0/1 | Solved |
| *IFT88* | ONRD | RP | NM_001318491.2: c.538G>T<br>(p. Val180Phe);<br>pathogenic | - | Monoallelic 0/1 | With evidence |
| *KIAA2022* | NRD | MD | NM_001008537.3: c.4385del<br>(p. Cys1462LeufsTer24);<br>likely pathogenic | - | Monoallelic 0/1 | With evidence |
| *TTPA* | IRD | RP | NM_000370.3: c.227_235del<br>(p. Trp76Ter);<br>likely pathogenic | - | Monoallelic 0/1 | With evidence |
| *TTPA* | IRD | MD | NM_000370.3: c.227_235del<br>(p. Trp76Ter);<br>likely pathogenic | - | Monoallelic 0/1 | With evidence |
| *CDHR1* | IRD | RP | NM_033100.4: c.2410_2485del<br>(p. Thr804ProfsTer12);<br>likely pathogenic | - | Monoallelic 0/1 | Partially solved |
| *CDHR1* | IRD | RP | NM_033100.4: c.2410_2485del<br>(p. Thr804ProfsTer12);<br>likely pathogenic | NM_033100.4: c.476C>A<br>(p. Ala159Glu)<br>(VUS) | Biallelic 0/1, 0/1 | Partially solved |

Another direct application of the IRD relative variant frequencies is the reevaluation of the clinical significance of VUS. We extracted the FC of IRD-AF compared to PC-AF for a set of manually curated VUS whose reclassification could contribute to a conclusive diagnosis of an IRD case in our cohort, and present in the final dataset (N=63). Of them, six VUS are IRD-MFVs (log2(FC)>=2.48, see Methods), and 11 VUS are more frequent in IRD cases than in PCs with a log2(FC)FC>=1.5. ACMG classification was performed for the 11 VUS adding the ACMG criteria PS4 (“The prevalence of the variant in affected individuals is significantly increased compared with the prevalence in controls”) as true. Of them, 10 (91%) were reclassified as likely pathogenic/pathogenic (Table 2). This analysis was able to solve two cases: one with the variant in dominant gene *COL11A1*, and one with the variant in *OFD1* that has X-linked inheritance (Table 2).

**Table 2.** Reclassification of VUS adding information about the difference in their frequency in cases and controls. The ACMG classification of VUS with log2 Fold change (FC) higher than 1,5 is reevaluated. Gene, variants in nucleotide and protein code are provided. The inheritance mode is annotated as autosomal recessive (AR), autosomal dominant (AD) and x-linked (XL). In “Varsome” column we add the automatic ACMG classification of the variants provide by the Varsome database at the time of writing. Previous ACMG criteria fulfilled by the cases are annotated in the “Criteria” column. Column “PS4” provides the new ACMG classification reached after including PS4 criteria. In column “Status” reclassified variants are marked.

| Gene | HGVSc | HGVSp | Inheritance | $\log_2(\text{FC})$ | VarSome | Criteria | PS4 | Status |
| --- | --- | --- | --- | --- | --- | --- | --- | --- |
| *CACNA1F* | NM_001256789.3:<br>c.4009-3C>G | - | XL | 2,7 | 3 | PM2, BP4 | 4 | Reclassified |
| *CDH23* | NM_022124.5:<br>c.4231G>A | p. Glu1411Lys | AR | 2,0 | 3 | PM2, PP3 | 4 | Reclassified |
| *CDHR1* | NM_001171971.3:<br>c.1589C>G | p. Thr530Ser | AR | 2,6 | 3 | PM1, PM2, BP1 | 4 | Reclassified |
| *COL11A1* | NM_080629.2:<br>c.4838C>A | p. Thr1613Asn | AD | 1,6 | 3 | PM2, PP2 | 4 | Reclassified |
| *GDF6* | NM_001001557.4:<br>c.125G>T | p. Gly42Val | AR/AD | 2,0 | 3 | PM1, PM2, PP5, BP6 | 4 | Reclassified |
| *IMPG2* | NM_016247.4:<br>c.1460A>T | p. His487Leu | AR/AD | 2,7 | 3 | PM2, BP4 | 4 | Reclassified |
| *MERTK* | NM_006343.3:<br>c.2435A>G | p. Tyr812Cys | AR | 2,0 | 3 | PM1, PM2, PP3, BP6 | 4 | Reclassified |
| *NYX* | NM_022567.2:<br>c.505A>G | p. Asn169Asp | XL | 2,9 | 3 | PM2 | 3 | Not reclassified |
| *OFD1* | NM_003611.2:<br>c.87T>G | p. Asp29Glu | XL | 2,7 | 3 | PM2, PP3, BP1 | 4 | Reclassified |
| *RP1* | NM_006269.2:<br>c.2497T>C | p. Phe833Leu | AR/AD | 1,7 | 3 | PM2, BP4 | 4 | Reclassified |
| *WFS1* | NM_001145853.1:<br>c.1597C>T | p. Pro533Ser | AR/AD | 3,7 | 3;4 | PM2, PP3 | 4 | Reclassified |

### 2.3. Prioritization of candidate genes based on weighted cohort-specific frequency of pathogenic and benign variants in non-solved IRD cases

In order to detect genes with an accumulated high pathogenicity in solved and non-solved IRD cases as good candidates to be involved in IRD phenotypes, for each IRD subcohort, we extracted deleterious and benign variants, and calculated the FC for their AF compared to the AF in PCs. For every subcohort and gene, the distributions of log2(FCs) in deleterious and benign variants were compared using a Wilcoxon rank sum test. Genes with a significant higher frequency in IRD cases of deleterious variants compared to benign variants (p-value<0.05) were selected. This revealed a number of genes with an accumulated pathogenicity in solved and non-solved IRD cases that we classified, as before, in three groups: IRD-genes, OERD-genes and NRD-genes. Actionable genes defined by ACMG were removed from the analysis (Supplementary Table S5). Thus, in IRD solved cases, we found 56 genes enriched in deleterious mutations. Of them, 22 (39%) were IRD genes, being the top 5 genes with more deleterious variants *ABCA4, USH2A, MYO7A, EYS* and *ADGRV1*, (Supplementary Table S6, and Figure 3A). In addition, 24 (43%) were OERD genes, highlighting here the top 5 with more deleterious variants *NEB, PAH, DNAH11, DNAH5* and *ATM* (Supplementary Table S6 and Figure 3A), and 10 (18%) were NRD genes, including top 5 genes *OBSCN, DYSF, SPTBN5, OTOF* and *SPTB*. Regarding non-solved cases we found 18 genes with an accumulated pathogenicity, with IRD-genes less represented (22%) and OERD-genes more present (55%) than in solved cases. Finally, 22% of the prioritized genes were NRD-genes (Figure 3B, and Table 3). Interestingly, there was a high overlap between IRD-genes and OERD-genes prioritized in solved and non-solved cases, 75% and 70% of the smallest group (non-solved cases), respectively. However, the overlap in NRD-genes was smaller, with only one gene (25% of genes in non-solved cases) found in both IRD subcohorts (Supplementary Figure S4).

**Table 3.** Genes having more frequent deleterious variants in IRD non-solved cases than in pseudocontrols. The genes are sorted by the gene panel where they are present (IRD, OERD and NRD) and by number of deleterious variants. The gene panel column refers to the type of diseases that the gene has been associated with, inherited retinal dystrophies (IRD), other eye related diseases (OERD) and other non-eye related diseases (NRD). Number of deleterious and benign variants in IRD non-solved cases are shown. FDR stands for False Discovery Rate of the Wilcoxon rank sum test performed. Inheritance mode associated for each gene is annotated in the “Inheritance” column, as recessive, dominant or R / D (either recessive or dominant).

| Gene | Deleterious | Benign | FDR | Gene Panel | Inheritance |
| --- | --- | --- | --- | --- | --- |
| *MYO7A* | 10 | 42 | 1.08E-03 | IRD | Recessive |
| *DYNC2H1* | 9 | 43 | 9.81E-04 | IRD | Recessive |
| *LAMA1* | 7 | 22 | 3.56E-02 | IRD | Recessive |
| *HMCN1* | 6 | 43 | 2.75E-02 | IRD | Dominant |
| *NEB* | 12 | 59 | 4.55E-03 | OERD | Recessive |
| *PAH* | 11 | 12 | 1.32E-02 | OERD | Recessive |
| *ALS2* | 8 | 11 | 1.87E-02 | OERD | Recessive |
| *DNAH9* | 7 | 13 | 1.32E-02 | OERD | Recessive |
| *HSPG2* | 6 | 44 | 1.87E-02 | OERD | R / D |
| *DNAH5* | 6 | 43 | 1.87E-02 | OERD | Recessive |
| *PLEC* | 6 | 80 | 1.87E-02 | OERD | Dominant |
| *ADAMTSL4* | 5 | 16 | 2.70E-02 | OERD | Recessive |
| *NDUFV1* | 5 | 8 | 1.32E-02 | OERD | Recessive |
| *COL4A3* | 5 | 15 | 1.87E-02 | OERD | R / D |
| *OBSCN* | 9 | 91 | 1.32E-02 | NRD | Recessive |
| *CAPN3* | 7 | 7 | 1.28E-02 | NRD | Recessive |
| *MYOM1* | 6 | 22 | 1.32E-02 | NRD | Recessive |
| *ABCB11* | 6 | 7 | 1.87E-02 | NRD | Recessive |

**Figure 3.**
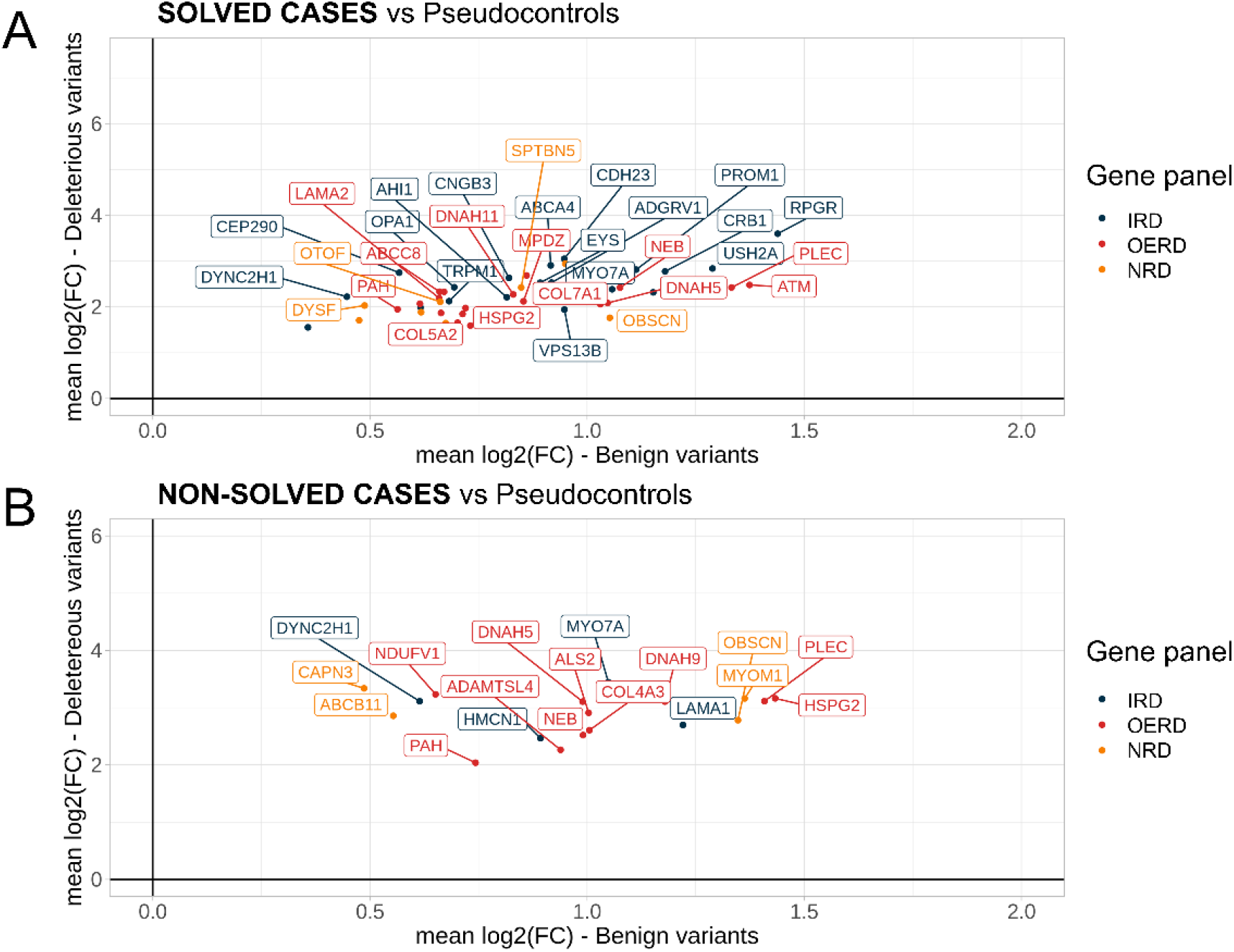
Genes with a higher accumulated pathogenicity in solved and non-solved IRD cases compared to pseudocontrols. Mean fold changes (FCs) in log2 scale for deleterious (Y-axis) and benign variants (X-axis) are shown for each gene. For IRD solved cases, significative genes with at least 8 deleterious variants are shown in the plot (A). For IRD non-solved cases all significative genes are shown (B).

In a reevaluation of the non-solved cases with pathogenic, likely pathogenic or VUS variants in the prioritized genes, we found five cases carrying mutations possibly associated to the phenotype in two IRD associated genes, and one OERD gene (Table 4). Variants found in the gene *MYO7A* explained this case phenotype and helped to fully characterize it. Regarding OERD genes, the two pathogenic variants found in the gene *ADAMTSL4* helped to solve lens luxation phenotype of this case.

**Table 4.** New diagnostic status produced by the reassessment of non-solved IRD cases with deleterious variants in the genes prioritized. Details about five IRD non-solved cases with variants adding knowledge to the phenotype. The gene panel column refers to the type of diseases that the gene has been associated with, inherited retinal dystrophies (IRD) and other eye related diseases (OERD). In the phenotype, abbreviations are MD: macular dystrophy. In variant column, HGVSc, HGVSp and ACMG classification are included. Genotype column has information about the genotype of the variant(s) in the sample. Diagnostic status column has the new diagnostic status of the case after the reanalysis, as: 1) “solved”, when the case has a conclusive diagnosis thanks to the variant(s) identified, or 2) “non-solved”, in any other case. Non-solved cases were annotated according to findings that may help in a future diagnosis as: 2.1) “partially solved”, if a heterozygous pathogenic or likely pathogenic variant is found within a recessive gene that fits the observed phenotype. And 2.2) “with evidence”, when the variants identified in the analysis are: 2.2.1) pathogenic/likely pathogenic variants in genes not yet associated with the disease but with some evidence published in the literature or with overlapping phenotypes, or 2.2.2) VUS in a gene associated to the phenotype when one VUS in dominant or 1-2 VUS in recessive genes were found.

| Gene | Gene panel | Phenotype | Variant (HGVS<sub>c</sub>, HGVS<sub>p</sub>, ACMG classification) | Genotype | Diagnostic status |
| --- | --- | --- | --- | --- | --- |
| *DYNC2H1* | IRD | MD | NM_001080463.2: c.3793C>T (p. Arg1265Cys), VUS;<br>NM_001080463.2: c.1468C>T (p. Arg490Cys), VUS | Biallelic 0/1, 0/1 | With evidence |
| *DYNC2H1* | IRD | MD | NM_001080463.2: c.988C>T p. Arg330Cys, pathogenic | Monoallelic 0/1 | Partially solved |
| *DYNC2H1* | IRD | MD | NM_001080463.2: c.7966C>T p. Arg2656Cys, likely pathogenic | Monoallelic 0/1 | Partially solved |
| *MYO7A* | IRD | Usher syndrome | NM_000260.4: c.1996C>T (p. Arg666Ter), pathogenic;<br>NM_000260.4: c.3764del (p. Lys1255ArgfsTer8), pathogenic | Biallelic 0/1,0/1 | Solved |
| *ADAMTSL4* | OERD | Lens luxation | NM_001288607.2: c.2594G>A (p. Ser865Asn), pathogenic;<br>NM_001288607.2: c.767_786del (p. Gln256ProfsTer3), pathogenic | Biallelic 0/1,0/1 | Solved |

### 2.4 Carrier’s frequency of recessive diseases from a multi-disease cohort

We calculated the carrier frequency (CF) for 69 genes involved in recessive non-syndromic IRDs using the frequency of pathogenic heterozygous variants located in IRD genes in the pseudocontrol subcohort. We found three genes with a CF >= 1%, which represents a total of ∼4% of the total analyzed (Supplementary Table S12). We highlight *ABCA4* and *USH2A* with a CF of ∼7% and ∼3% respectively (Figure 4), the first being responsible of Stargardt disease, and the later causing Usher syndrome. In the case of ABCA4 if hypomorphic variants are excluded as in a previous work performed by Hanany and collaborators [19], CF is reduced to ∼4%. These genes are also the two most frequent in our IRD subcohort, found causal in 21% and 15% of the cases respectively (data not shown). The most frequent variant for *ABCA4* in the IRD subcohort and PC subcohort were NM_000350.3: c.3386G>T and NM_000350.3: c.3113C>T respectively, while *USH2A* had variant NM_007123.5: c.2276G>T as the most frequent in the two subcohorts. The CF calculated using deleterious heterozygous/hemizygous variants on dominant/X-linked genes is available in Table S14.

**Figure 4.**
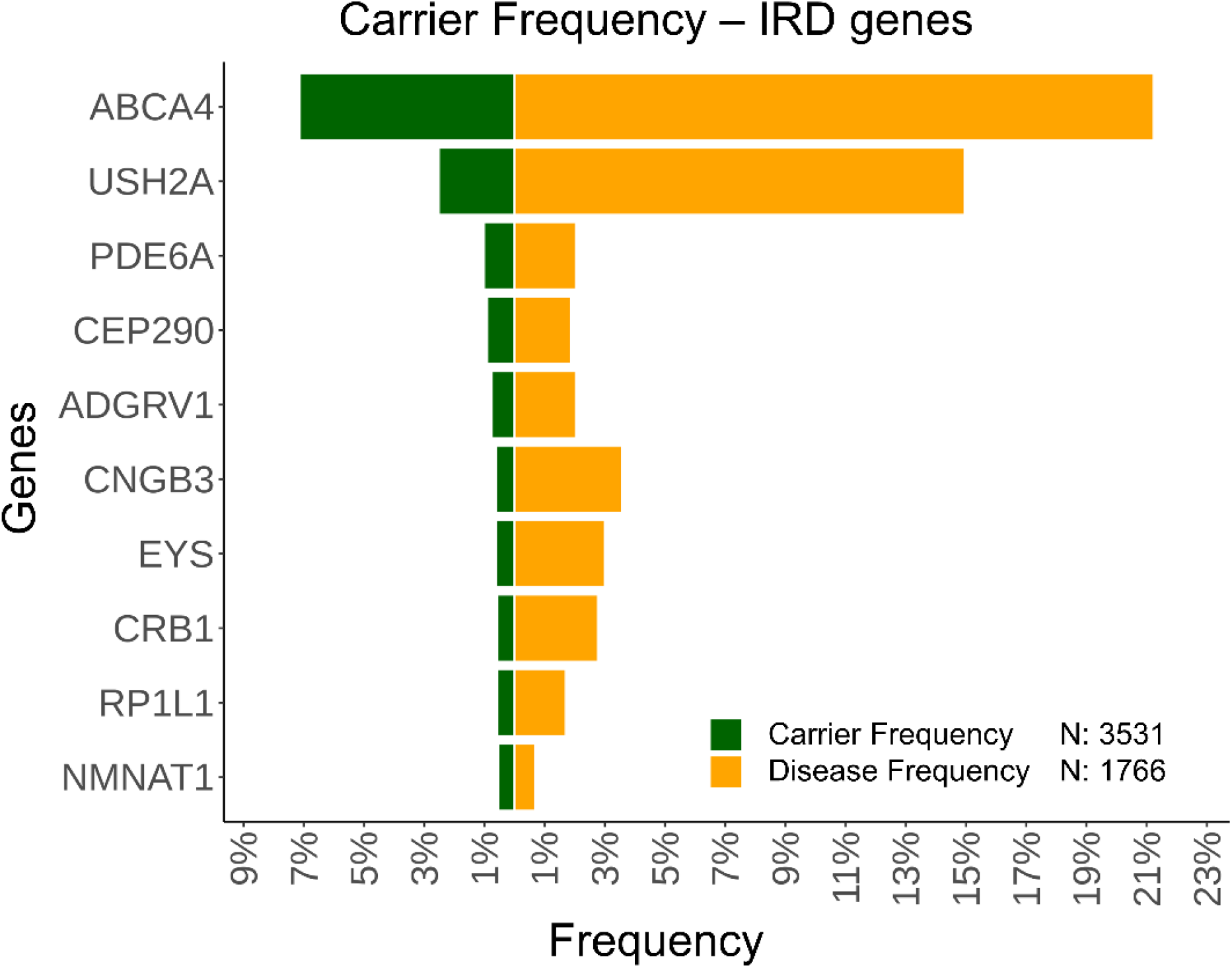
Carrier frequency of pathogenic variants in recessive IRD genes in pseudocontrols. The carrier frequency (CF) was calculated for all IRD genes with a recessive inheritance pattern, and at least one solved case in our cohort. In green is represented the CF of the genes and in orange the frequency in our IRD subcohort. The pseudocontrol subcohort was composed of 3531 cases, and the IRD subcohort had 1766 cases. Top 10 genes with higher CF are shown ordered decreasingly.

## 3. Discussion

The recent application of NGS techniques has considerably increased our competence to study and diagnose rare diseases. Regarding IRDs, although new associated genes are still being discovered (see RetNet, https://sph.uth.edu/retnet), the diagnostic ratios need to be boosted, since up to ∼46% remain unsolved (data from our own IRD cohort at the time of writing). At the same time, at the diagnostic setting, the genomic information from patients is being accumulated as databases or annotation systems, that can also be privative if only commercial solutions are used. Initiatives like gnomAD [17] or the Collaborative Spanish Variability Server [18] are acting as a crowdsource to recover this data from and to the community and offer it back as aggregated allele frequencies. Allelic frequency calculations have been proven to be useful in studying the prevalence of rare diseases [50], or integrated in the diagnostic analysis routines, as an additional annotation source to detect technical biases [51], or to interpret and classify variants [52]. These resources have also been used to study the CF of deleterious variants in recessive genes in a particular population [16]. In this study, we propose to develop a framework of methods able to prioritize variants and genes in order to explore new associations with a specific disease. Thus, the reuse of the genomic data may provide new discovery capabilities to a heterogeneous cohort of genetic diseases, an extra value to the resources invested previously, as well as allowing the patients to contribute to their own or others’ future diagnosis. The main hypothesis behind this work is that, aside from a few causing mutations, the genome of patients with Mendelian diseases behaves similarly as those of healthy population, so patients with non-related diseases can act as pseudocontrols between each other. The fact of focusing on a heterogeneous but single-center cohort has two main advantages against using controls from larger genomic population databases [12,13,17]: first they can be technically more similar to the sequencing produced on the patients of interest; second, the geographical origin bias can be better controlled [15]; last, the phenotyping of patients can be fine-tuned by experts in order to create subcohorts of interest. A concept introduced here is the significantly different frequent variants, named as MFVs, that are variants more present in cases than in pseudocontrols, with statistical support. As a proof of concept, we found that considering the whole IRD cohort, the IRD-MFVs are enriched in deleterious mutations, suggesting that the prioritization is effective there. Indeed, variants selected on solved IRD cases in IRD genes are mostly pathogenic (>78%) compared to benign, due to the detection of prevalent causal variants. In contrast, non-solved IRD cases show no differences in the proportion of deleterious / benign in prioritized and non-prioritized variants in IRD related genes, indicating that there are not many described pathogenic variants left out during diagnosis. In spite of this, our approach was able to solve or partially solve five cases with prioritized variants in IRD genes. We also highlight here the significant accumulation of deleterious variants in those prioritized located in OERD and NRD genes, in both IRD solved and non-solved cases. The rationale behind it might be different, though. While frequent pathogenic mutations in OERD and NRD genes in solved cases may suggest a more complex genotype scenario for Mendelian diseases [53,54], in non-solved cases we have to add the possibility of needing a disease re-evaluation, and the causing mutation being present in a not yet associated IRD gene. The exploration of syndromic and non-syndromic forms separately provides the same signal but with lower p-values for syndromic cases. Up to 8 non-solved cases gained new insights due to the reevaluation of our prioritized variants. Our variant prioritization approach was also applied for VUS reclassification, a major challenge to unlock the diagnosis of rare pathologies [10,55–57]. Indeed, in our IRD subcohort, top 5 genes with more deleterious variants in IRD solved cases, present a higher degree of uncertainty in variant annotation (proportion of VUS and deleterious variants) in unsolved IRD cases. For an initial list of 63 VUS whose reclassification may solve a case from the IRD cohort, we found 11 VUS more frequent in IRD cases than in pseudocontrols and 10 of them (∼91%) changed their classification to likely pathogenic or pathogenic by the application of the ACMG PS4 criteria. This reclassification was able to solve two cases and provide new evidence to other 10 patients.

In addition to the revised cases reported in this work, the disease specific AF and its comparison to pseudocontrols have been implemented in the variant annotation task of our reanalysis pipeline [34] that performs periodic reanalysis of non-solved cases as well as WES and WGS analysis. Thus, the database is expected to contribute to the diagnosis of more patients over time. Furthermore, and in order to facilitate the implementation of the database and adjust it to the cohorts of other clinical settings, we have code and instructions to build an in-house database available at https://github.com/TBLabFJD/DbofAFs.

In parallel, we also aimed to highlight genes besides variants. Our method extracts genes having more frequent deleterious variants in IRD cases than in pseudocontrols, weighted by the relative frequency of benign variants in order to increase the disease association signal. Several discovery scenarios may fit into the results of this proposal. First, finding underrated IRD genes with a role in IRD cases, such as *DYNC2H1* and *MYO7A* genes that may carry not previously inspected pathogenic variants. Thus, we were able provide new evidence in four cases. Next, providing extra findings in complex cases, either syndromic cases, gene modifiers, or dual diagnosis. For instance, we have a dual diagnosis in a previously non-solved syndromic IRD case with rod-cone dystrophy (HP:0000510) and lens luxation (HP:0012019) among other systemic findings. This case has a partially solved IRD phenotype with a heterozygous variant in gene *CDH23* (Table 1), and a solved Lens Luxation phenotype with two pathogenic variants in compound heterozygosis in gene *ADAMTSL4*. In non-classical Mendelian scenarios, the exploration of the mutational landscape of IRD may help for instance to identify more complex cases as: i) genetic pleiotropy together with causal variants in recessive forms [22], ii) digenic inheritance [23], or iii) triallelic sites associated to BBS [24]. Last, there is also the possibility of detecting genes not yet associated to IRD, that is, candidate genes that may become IRD genes if further analyses are performed.

As result of the reevaluation of IRD non-solved cases using the methods described in this work, we were able to solved four cases (one of them with a dual diagnosis still unsolved), and provide new insights in 20 cases, 15 of them providing a single variant for a recessive case that fits the phenotype (partially solved), and five with candidate variants in genes not yet fully associated to the phenotype (marked here as “with evidence”). Further studies are needed to solve cases annotated as partially solved and with evidence.

An additional interesting use of an internal database of allele frequencies is to have a cohort-specific CF estimation. This analysis can provide a better understanding on how deleterious variants are distributed in a general population and is relevant for their use in a public health strategy for genetic counselling. For instance, in the case of IRD, the gene with a higher carrier frequency is *ABCA4*, with carrier variants in ∼7% of the population, which is in line with previous estimations [58]. Considering the curated set of pathogenic variants used by Hanany et al [19] for *ABCA4*, we obtained a similar CF (∼6%). Nevertheless, excluding hypomorphic variants, as recommended in this study, CF drops to ∼4%. The high CF obtained for this gene in our cohort can be explained partially by these variants.

It is reasonable to state that although pseudocontrols are suitable for providing a good estimation of general allele frequencies, the lack of healthy controls can be seen as a limitation. However, availing of such control sample set is not always feasible for clinical setting and should be provided under the umbrella of national plans. The major constraint in the discovery capability of our database is that we are restricted to the ∼5000 genes targeted in the clinical exome approaches, and thus, an implementation using data from whole exomes would be optimal. Our intention is to maintain and expand the database in number of cases but also in genomic regions. We should mention that IRD non-solved cases presented in the cohort can also have causing variants in non-coding regions, which are not covered with the clinical exome approach.

Although in this work we focus on IRD as the larger group of diseases in our cohort, the same methodology can be applied to other genetic rare diseases in our cohort as well as in other settings. In addition, the disease-specific subcohort that is subject of analysis can also be tuned in its composition in order to provide different capabilities to the discovery process. For instance, although our subcohort of IRD unsolved cases did not include suspected recessive cases with only one detected candidate variant in heterozygosity (the so-called monoallelic cases), they could be included in this discovery subcohort so these candidate variants are also evaluated.

In conclusion, our cohort-specific database of allele frequencies has proven to be able to diagnose non-solved IRD cases, reclassify VUS, propose candidate genes, and calculate CF on genes of interest. We believe that the results shown here can highlight the importance of the reuse of genomic data produced in clinical settings, where the phenotyping is usually exhaustive and the patients waiting for a diagnosis or a genetic counselling can be directly benefited.

## 4. Materials and Methods

### 4.1 Ethics Approval and consent to participate

The project was reviewed and approved by the Research Ethics Committee of UH-FJD (Ref. 2016/ 59) and fulfills the principles of the Declaration of Helsinki and subsequent reviews. All patients signed an informed consent before participating. All samples included in this work were pseudonymized and genomic data was only treated in aggregation.

### 4.2 Cohort description

We retrospectively selected all index cases (N=5683) with a clinical exome test performed as a first-tier approach at the Genetics and Genomics Department of the University Hospital Fundación Jiménez Díaz (UH-FJD, Madrid, Spain) from September-2015 to May-2021. The cohort included patients suffering from genetic diseases classified in 14 categories, with the largest disease group being IRD (N=1766) (Supplementary Table S7). The rest of the diseases were grouped in “other eye related diseases” (OERD) and “non-related diseases” (NRD). Based on the diagnostic status set by the molecular geneticists after CE inspection, all IRD cases were classified as solved and non-solved. From the non-solved cases, we extract those annotated as “partially solved” or “VUS-cases”.

### 4.3 Sequencing tests

Samples were analyzed using targeted DNA sequencing with two different commercial sequencing panels: TruSightOne Sequencing Panel kit (TSO, Illumina, San Diego, CA), and Clinical Exome Solution Sequencing Panel kit (CES, Sophia Genetics, Boston, MA). CES panel targets a total of 4828 genes and regulatory regions and TSO targets 4813 genes, with an overlap of 3567 genes between both panels (Supplementary Figure S5).

### 4.4 Bioinformatics reanalysis

In order to have a homogeneous variant calling and annotation of all sequencing tests, all sequenced data was reanalyzed using a custom bioinformatics pipeline for both single nucleotide variants (SNVs) and small insertions and deletions (indels) [25]. This pipeline included exonic, intronic, and UTR analysis. For variant calling, we included 1000 base pairs padding for each target region, for both, TSO and CES clinical exome tests. The pipeline is based on GATK 4.1 variant caller [26], and uses BWA-MEM aligner [27] to the GRCh37/hg19 reference genome. The following databases were used for annotation: i) allele frequency: gnomAD [17], 1000genomes [28], and Kaviar [29]; ii) pathogenicity prediction: SIFT [30], PolyPhen [31], CADD [32], LRT [33], M-CAP [34], MetaLR [35], MetaSVM [35], MutationAssesor [36], MutationTaster [37], PROVEAN [38], and FATHMM [39]; iii) splicing prediction: ada_score [40] and rf_score [40]; iv) ClinVar [41]; v) conservation: phastCons20way [42] and phyloP20way [43]; vi) gene tolerance to loss of function (LoF) variants: LoFtool [44], and ExACpLI [12]; vii) constrained coding regions by means of gnomAD_CCR [45]; and viii) potential loss of heterozygosity regions, annotated with PLINK [46]. The pipeline is available at https://github.com/TBLabFJD/VariantCallingFJD.

### 4.5 Detection and removal of sample duplicates and cryptic relatedness

All known sample duplicates and relatives were removed prior to frequency calculation. In order to detect other possible sample duplicates and relatives, PLINK whole genome association analysis toolkit [46] was used to calculate inbreeding coefficients (identity--by-descent, IBD). First, single nucleotide polymorphisms (SNPs) pruning was performed removing SNPs covered in less than 95% of the samples (PLINK parameter: geno 0.05), with less than 5% allelic frequency (PLINK parameter: maf 0.05), and in linkage disequilibrium (PLINK parameters: indep-pairwise 50 5 0.5). With the resulting SNPs, IBD was calculated for all sample combinations (PLINK parameter: genome). All samples with a PI_HAT score higher than 0.35 were removed.

### 4.6 Variant frequency calculation for IRD patients and pseudocontrols

After identifying and removing sample duplicates and relatedness (N=5683), variants (in vcf-format) from the index-cases were processed together and merged into a multi-vcf file. Sequencing coverage was also calculated for each sample to distinguish between non-covered and non-mutated sites.

We developed in-house routines for allelic frequency calculation based on Hail python library for genomics data exploration and analysis (https://hail.is). The allele frequency (AF), allele-number (AN), allele-count (AC) and homozygotes-count were obtained for the general cohort and for several subcohorts composed of IRD cases: 1) all IRD cases, 2) non-solved IRD cases, 3) solved-IRD cases, 4) syndromic-IRD cases. 5) non-syndromic IRD cases, and 6) macular dystrophies cases. To define the subcohort of IRD PC for all IRD subcohorts, we took samples from the subcohort NRD (N=3531). AF, AN and AC were calculated from this PC subcohort (PC-AF, PC-AN and PC-AC).

### 4.7 Definition of genes associated to IRD, OERD and NRD

Three disease specific gene panels were used in the inspection of variants and genes: i) IRD gene panel (244 genes; including 136 genes for syndromic-IRD and 108 genes for non-syndromic IRD) as the virtual gene panel used in the diagnosis of IRD cases in the Genetics and Genomics Department of the UH-FJD, and extracted using RetNet, HGMD and literature searches (Supplementary Table S8), ii) OERD genes, including non-IRD genes with ocular phenotype (all genes linked with the HPO term “Eye Disease”–HP:0000478, N=1542 genes, Supplementary Table S9), and iii) NRD genes (the rest of the genes included in TSO/CES panels, not related with eye diseases, N=3260, Supplementary Table S10). Genes which are recommended by ACMG to report in case of secondary findings [47] (Supplementary Table S5) were excluded from the gene panels and analyses but the gene *RPE65* that belongs to the IRD panel.

### 4.8 Variants discarded for analysis

Variants detected in the 5683 samples from our general cohort were further filtered out using two criteria: i) quality filtering, we removed 5% of variants with lowest AN, and ii) population filtering, in order to discard a population origin bias in our IRD subcohort compared to the rest of the cohort, we keep variants present in IRD solved or non-solved cases, and assuming no differences in population origin between IRD solved and non-solved cases, having a fold change between non-solved-AF and solved-AF >90% percentile, from them we rescue those having a non-solved-AF and solved-AF <0.1.

### 4.9 Determination of differentially frequent variants in IRD subcohorts compared to pseudocontrols

We define differentially frequent variants as those that have a higher frequency in a subcohort compared with a control subcohort. In order to extract variants differentially frequent in the IRD subcohorts (solved-IRD, non-solved IRD, syndromic-IRD, non-syndromic IRD and macular dystrophies) compared to the IRD PC subcohort, we calculated the FC of the AF in the IRD subcohort compared to the PC-AF for each of the variants. Based on the distribution of the log2 of FCs (log2(FC)) of all variants, we selected those above the 90% as the significant differential frequent variants in a subcohort, tagged as IRD-MFV (IRD most frequent variants) for any IRD subcohort. Variants annotated by ClinVar as “pathogenic” or “likely pathogenic” or with a CADD_PHRED≥30 (top 0.1% most deleterious variants according to CADD) were classified as deleterious, and variants annotated by ClinVar as “benign” or “likely benign” were classified as benign. We compared the proportion of deleterious variants in the IRD-MFV group with the rest of the variants (non-prioritized variants) for solved-IRD and non-solved-IRD cases. Furthermore, we performed this comparison grouping IRD cases as: syndromic forms, non-syndromic forms and macular dystrophies. A Fisher’s exact-test was applied to compare the proportion of deleterious variants in these groups, p-value<0.05 was taken as significant.

### 4.10 VUS reclassification

We selected VUS whose reclassification can determine the diagnosis of an IRD case in our cohort. These VUS are reported in the diagnostic process at the Genetics Department of the UH-FJD if no pathogenic or likely-pathogenic variant are found associated to the phenotype. Variants are classified using ACMG guides. In the IRD subcohort, there were reported 100 VUS fulfilling these criteria [10] (Supplementary Table S11). Of these, 63 fulfilled criteria to be within the database generated in this work, and were still classified as VUS according to ACMG (information taken from VarSome at the time of the analysis). For these VUS, we annotated IRD-AF (general IRD cohort) and PC-AF frequencies, calculated the FC for these two frequencies and selected two sets: 1) VUS with a log2(FC)>=1.5 (N=11), VUS with a log2(FC)>=2.48 (value of the 90 percentile of the distributions of the log2(FC), N=6). For all selected VUS (N=11) we marked the specific ACMG criterion PS4 for which (“The prevalence of the variant in affected individuals is significantly increased compared with the prevalence in controls”) and applied previous evidences to obtain a new ACMG classification.

### 4.11 Gene prioritization for IRD association

To prioritize genes in non-solved IRD cases, we selected for each gene included in the database: i) deleterious variants annotated by ClinVar as “pathogenic” or “likely-pathogenic” or with a CADD_PHRED≥30, and ii) benign variants annotated by ClinVar as “benign” or “likely benign”. Genes with at least five deleterious and five benign variants were selected for further analysis. For each selected variant, a log2(FC) was calculated between non-solved IRD-AF and PC-AF. Finally, we applied Wilcoxon rank sum test to the distribution of log2(FC) for deleterious and benign variants in each gene. P-values were adjusted using FDR and genes with an adjusted p-value<0.05 were considered significant. This list of significant genes was classified into three different gene panels according to the relation degree with IRDs: i) IRD gene panel, ii) OERD gene panel, and iii) NRD gene panel. This analysis was also performed for solved-IRD cases. (Supplementary Figure S6)

### 4.12 Carrier frequency calculation

Carrier frequency (CF) was calculated for genes in the non-syndromic IRD gene panel with at least three solved cases in our cohort. Genes were classified as with autosomal recessive or dominant inheritance patterns using the software DOMINO [48] and OMIM database [49]. In genes annotated as recessive, CF was calculated including variants classified: i) “pathogenic” or “likely pathogenic” in ClinVar; ii) or “pathogenic” or “likely pathogenic” in LOVD database; iii) or with a CADD_PHRED score ≥30; iv) or frameshift/stop-gain variants. The total AC of the variants selected was divided by the AN and the result multiplied by 2 (two alleles) and multiplied by 100 to represent the result as a percentage (0-100%), according to equation 1.

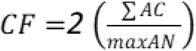

**Equation 1**. Carrier frequency (CF) calculation.

For the gene *ABCA4*, the CF was also calculated excluding hypomorphic variants as described in Hanany et al [19].

## Data Availability

All data produced in the present study are available upon reasonable request to the authors

