## Supplementary for "Aggregated genomic data as cohort-specific allelic frequencies can boost variants and genes prioritization in non-solved cases of inherited retinal dystrophies"

### Supplementary Tables

**Table S1.** Number of deleterious and benign within the prioritized (IRD-MFV) and non-prioritized sets in IRD solved cases, including those in all genes and in genes from the inherited retinal dystrophies (IRD) panel, the other eye related diseases (OERD) and non-eye related diseases (NRD). P-values of the Fishers' exact test are shown.

| | GENE_PANEL | Clinical Significance | IRD-MFV | Non-prioritized | p-value |
|---|---|---|---|---|---|
| SOLVED | ALL genes | Deleterious | 404 | 4191 | 4.77E-56 |
| | | Benign | 762 | 23020 | |
| | IRD genes | Deleterious | 177 | 593 | 5.97E-61 |
| | | Benign | 49 | 1926 | |
| | OERD genes | Deleterious | 80 | 1457 | 1.71E-06 |
| | | Benign | 314 | 10843 | |
| | NRD genes | Deleterious | 147 | 2141 | 2.75E-08 |
| | | Benign | 399 | 10251 | |

**Table S2.** Number of deleterious and benign within the prioritized (IRD-MFV) and non-prioritized sets in IRD non-solved cases, including those in all genes and in genes from the inherited retinal dystrophies (IRD) panel, the other eye related diseases (OERD) and non-eye related diseases (NRD). P—values of the Fishers' exact test are shown.

| | GENE_PANEL | Clinical Significance | IRD-MFV | Non-prioritized | p-value |
|---|---|---|---|---|---|
| NON-SOLVED | ALL genes | Deleterious | 148 | 2143 | 1.69E-32 |
| | | Benign | 399 | 21222 | |
| | IRD genes | Deleterious | 7 | 193 | 6.6E-02 |
| | | Benign | 31 | 1844 | |
| | OERD genes | Deleterious | 37 | 770 | 3.25E-06 |
| | | Benign | 186 | 9805 | |
| | NRD genes | Deleterious | 104 | 1180 | 1.90E-28 |
| | | Benign | 182 | 9573 | |

**Table S3.** Number of deleterious and benign within the prioritized (IRD-MFV) and non-prioritized sets in IRD solved syndromic (SY), IRD non-syndromic (NSY) and macular dystrophies (MD) cases, including those in all genes and in genes from the inherited retinal dystrophies (IRD) panel, the other eye related diseases (OERD) and non-eye related diseases (NRD). P-values of the Fishers' exact test are shown.

| | GENE_PANEL | IRD type | Clinical Significance | IRD-MFV | Non-prioritized | p-value |
|---|---|---|---|---|---|---|
| SOLVED | IRD genes | SY | Benign | 11 | 427 | 7.84E-06 |
| | | | Deleterious | 13 | 68 | |
| | | NSY | Benign | 23 | 585 | 8.09E-26 |
| | | | Deleterious | 86 | 214 | |
| | | MD | Benign | 18 | 1099 | 2.19E-25 |
| | | | Deleterious | 56 | 239 | |
| | OERD genes | SY | Benign | 52 | 4184 | 2.29E-08 |
| | | | Deleterious | 19 | 257 | |
| | | NSY | Benign | 207 | 7094 | 2.28E-06 |
| | | | Deleterious | 52 | 801 | |
| | | MD | Benign | 153 | 6076 | 1.35E-04 |
| | | | Deleterious | 34 | 618 | |
| | NRD genes | SY | Benign | 68 | 4126 | 1.89E-12 |
| | | | Deleterious | 36 | 414 | |
| | | NSY | Benign | 228 | 6795 | 1.67E-13 |
| | | | Deleterious | 103 | 1176 | |
| | | MD | Benign | 200 | 5897 | 3.02E-03 |
| | | | Deleterious | 48 | 871 | |

**Table S4.** Number of deleterious and benign within the prioritized (IRD-MFV) and non-prioritized sets in IRD non-solved syndromic (SY), non-syndromic (NSY) and macular dystrophies (MD) cases, including those in all genes and in genes from the inherited retinal dystrophies (IRD) panel, the other eye related diseases (OERD) and non-eye related diseases (NRD). P-values of the Fishers' exact test are shown.

| | GENE_PANEL | IRD type | Clinical Significance | IRD-MFV | Non-prioritized | p-value |
|---|---|---|---|---|---|---|
| NON-SOLVED | IRD genes | SY | Benign | 7 | 355 | 1.39E-06 |
| | | | Deleterious | 7 | 14 | |
| | | NSY | Benign | 16 | 494 | 3.71E-04 |
| | | | Deleterious | 8 | 39 | |
| | | MD | Benign | 29 | 935 | 1.03E-01 |
| | | | Deleterious | 5 | 73 | |
| | OERD genes | SY | Benign | 57 | 3297 | 9.57E-18 |
| | | | Deleterious | 25 | 100 | |
| | | NSY | Benign | 145 | 5742 | 1.42E-06 |
| | | | Deleterious | 31 | 415 | |
| | | MD | Benign | 118 | 5176 | 7.03E-10 |
| | | | Deleterious | 31 | 311 | |
| | NRD genes | SY | Benign | 29 | 3350 | 2.69E-26 |
| | | | Deleterious | 35 | 194 | |
| | | NSY | Benign | 129 | 6795 | 7.84E-23 |
| | | | Deleterious | 76 | 1176 | |
| | | MD | Benign | 105 | 5186 | 4.57E-18 |
| | | | Deleterious | 54 | 499 | |

**Table S5.** Genes that ACMG recommends to report secondary findings. These genes are filtered out from the OERD and NRD gene panels.

| Gene | | | |
|---|---|---|---|
| ACTA2 | HFE | PCSK9 | TGFBR2 |
| ACTC1 | HNF1A | PKP2 | TMEM127 |
| ACVRL1 | KCNH2 | PMS2 | TMEM43 |
| APC | KCNQ1 | PRKAG2 | TNNI3 |
| APOB | LDLR | PTEN | TNNT2 |
| ATP7B | LMNA | RB1 | TP53 |
| BMPR1A | MAX | RET | TPM1 |
| BRCA1 | MEN1 | RPE65 | TRDN |
| BRCA2 | MLH1 | RYR1 | TSC1 |
| BTD | MSH2 | RYR2 | TSC2 |
| CACNA1S | MSH6 | SCN5A | TTN |
| CASQ2 | MUTYH | SDHAF2 | VHL |
| COL3A1 | MYBPC3 | SDHB | WT1 |
| DSC2 | MYH11 | SDHC | |
| DSP | MYH7 | SDHD | |
| ENG | MYL2 | SMAD3 | |
| FBN1 | MYL3 | SMAD4 | |
| FLNC | NF2 | STK11 | |
| GAA | OTC | TGFBR1 | |
| GLA | PALB2 | TGFBR2 | |

**Table S6.** Genes prioritized in solved cases of inherited retinal dystrophies. They are classified in three gene panels: genes from the inherited retinal dystrophies (IRD) panel, other eye related diseases (OERD) and non-eye related diseases (NRD).

| Gene | Deleterious | Benign | FDR | Gene Panel |
|---|---|---|---|---|
| ABCA4 | 84 | 24 | 1.13E-08 | IRD |
| USH2A | 73 | 76 | 7.35E-11 | IRD |
| MYO7A | 31 | 41 | 3.94E-06 | IRD |
| EYS | 20 | 11 | 3.20E-03 | IRD |
| ADGRV1 | 14 | 67 | 6.73E-04 | IRD |
| CRB1 | 13 | 7 | 4.81E-02 | IRD |
| PROM1 | 13 | 7 | 1.59E-02 | IRD |
| CNGB3 | 13 | 8 | 5.10E-03 | IRD |
| VPS13B | 13 | 37 | 1.59E-02 | IRD |
| CEP290 | 12 | 23 | 5.67E-04 | IRD |
| RPGR | 10 | 7 | 1.62E-02 | IRD |
| CDH23 | 9 | 35 | 1.77E-03 | IRD |
| DYNC2H1 | 9 | 39 | 7.28E-04 | IRD |
| TRPM1 | 9 | 14 | 1.92E-02 | IRD |

| | | | | |
|---|---|---|---|---|
| AHI1 | 9 | 29 | 2.41E-03 | IRD |
| OPA1 | 8 | 19 | 2.74E-03 | IRD |
| HMCN1 | 7 | 45 | 1.26E-02 | IRD |
| COL11A1 | 6 | 36 | 1.47E-02 | IRD |
| PRPF31 | 6 | 5 | 2.07E-02 | IRD |
| BBS9 | 6 | 9 | 1.52E-02 | IRD |
| RPE65 | 5 | 5 | 2.89E-02 | IRD |
| GRM6 | 5 | 14 | 2.24E-02 | IRD |
| NEB | 22 | 84 | 1.23E-05 | OERD |
| PAH | 18 | 8 | 1.38E-02 | OERD |
| DNAH11 | 14 | 36 | 1.70E-04 | OERD |
| DNAH5 | 13 | 47 | 1.22E-02 | OERD |
| ATM | 12 | 60 | 1.22E-02 | OERD |
| HSPG2 | 11 | 53 | 9.11E-04 | OERD |
| LAMA2 | 9 | 49 | 1.75E-03 | OERD |
| PLEC | 9 | 87 | 1.62E-02 | OERD |
| MPDZ | 9 | 28 | 6.33E-03 | OERD |
| ABCC8 | 8 | 9 | 1.26E-02 | OERD |
| COL5A2 | 8 | 36 | 4.81E-02 | OERD |
| COL7A1 | 8 | 26 | 3.67E-02 | OERD |
| C1QTNF5 | 7 | 8 | 1.59E-02 | OERD |
| DNAH9 | 7 | 6 | 1.44E-02 | OERD |
| FLNB | 7 | 37 | 1.78E-02 | OERD |
| DOCK8 | 7 | 28 | 1.62E-02 | OERD |
| CENPJ | 6 | 8 | 2.89E-02 | OERD |
| FANCI | 6 | 24 | 3.19E-02 | OERD |
| GALK1 | 6 | 9 | 1.28E-02 | OERD |
| XYLT1 | 5 | 11 | 1.42E-02 | OERD |
| ABCA12 | 5 | 15 | 4.04E-02 | OERD |
| COL4A3 | 5 | 15 | 2.31E-02 | OERD |
| MME | 5 | 5 | 4.57E-02 | OERD |
| EFHC1 | 5 | 14 | 1.52E-02 | OERD |
| OBSCN | 16 | 95 | 1.62E-02 | NRD |
| DYSF | 9 | 43 | 2.74E-03 | NRD |
| SPTBN5 | 8 | 8 | 3.05E-02 | NRD |
| OTOF | 8 | 24 | 6.49E-03 | NRD |

| | | | | |
|---|---|---|---|---|
| SPTB | 6 | 24 | 4.04E-02 | NRD |
| OBSL1 | 6 | 30 | 1.59E-02 | NRD |
| ANO3 | 5 | 13 | 1.59E-02 | NRD |
| ZFHX3 | 5 | 10 | 2.89E-02 | NRD |
| MYO1C | 5 | 16 | 2.37E-02 | NRD |
| CLCN1 | 5 | 21 | 2.24E-02 | NRD |

**Table S7**. Diseases included in the allele frequency database, and number (N) of cases of each disease. Diseases are classified in three categories: i) Inherited Retinal Dystrophies (IRD), ii) other eye related diseases (OERD) and iii) non-related diseases (NRD).

| Disease | N | Group |
|---|---|---|
| Inherited retinal dystrophies | 1766 | IRD |
| Mixed conditions | 1648 | NRD |
| Encephalopathies-ID-Epilepsy | 735 | NRD |
| Cardiopathy | 325 | NRD |
| Peripheral neuropathies | 209 | NRD |
| Optic atrophy | 200 | OERD |
| Hearing loss | 149 | NRD |
| Polymalformative syndromes | 123 | NRD |
| Neurodegeneration | 123 | NRD |
| Congenital eye defects | 107 | OERD |
| Nephropathy | 90 | NRD |
| Corneal Dystrophy | 80 | OERD |
| Metabolic | 65 | NRD |
| Myopathy | 63 | NRD |
| **TOTAL** | **5683** | |

**Table S8.** Gene panel used in the diagnosis of cases with inherited retinal dystrophies (IRD).

| Gene | | | | |
|---|---|---|---|---|
| ABCA4 | GUCY2D | RBP3 | BBS9 | NR2F1 |
| ABHD12 | HGSNAT | RBP4 | C5orf42 | OTX2 |
| ADAM9 | HK1 | RCBTB1 | CC2D2A | PANK2 |
| ADAMTS18 | HMCN1 | RD3 | CDH23 | PAX2 |
| ADGRV1 | IDH3B | RDH12 | CDH3 | PCDH15 |
| ADIPOR1 | IFT140 | RDH5 | CEP164 | PDZD7 |
| AFG3L2 | IFT172 | RGR | CEP41 | PEX1 |
| AHR | IMPDH1 | RGS9 | CIB2 | PEX2 |
| AIPL1 | IMPG2 | RGS9BP | CISD2 | PEX6 |
| ARL3 | ITM2B | RHO | CLN3 | PEX7 |
| ARL6 | KCNJ13 | RIMS1 | CLN5 | PGK1 |

| | | | | |
|---|---|---|---|---|
| ATF6 | KCNV2 | RLBP1 | CLN6 | PHYH |
| BBS1 | KLHL7 | ROM1 | CLN8 | PNPLA6 |
| BBS2 | LCA5 | RP1 | CNNM4 | POMGNT1 |
| BEST1 | LRAT | RP1L1 | COL11A1 | PPT1 |
| C2orf71 | LRIT3 | RP2 | COL11A2 | PRPS1 |
| C8orf37 | MAK | RP9 | COL2A1 | RPGRIP1L |
| CA4 | MERTK | RPE65 | COL9A1 | SDCCAG8 |
| CABP4 | MFRP | RPGR | COL9A2 | SLC41A1 |
| CACNA1F | MFSD8 | RPGRIP1 | COL9A3 | SLC9A6 |
| CACNA2D4 | MKKS | RS1 | CTSD | SPG7 |
| CDHR1 | MKS1 | SAG | DNAJC5 | TCTN1 |
| CEP290 | NDP | SEMA4A | DYNC2H1 | TCTN2 |
| CERKL | NEUROD1 | SLC24A1 | FLVCR1 | TIMM8A |
| CHM | NMNAT1 | SNRNP200 | GALE | TMEM138 |
| CLRN1 | NR2E3 | SPATA7 | GLIS2 | TMEM216 |
| CNGA1 | NRL | TEAD1 | GNPTG | TMEM237 |
| CNGA3 | NYX | TIMP3 | GRN | TMEM67 |
| CNGB1 | OAT | TMEM126A | HARS | TPP1 |
| CNGB3 | OFD1 | TOPORS | HMX1 | TREX1 |
| CRB1 | OPA1 | TRPM1 | IFT80 | TRIM32 |
| CRX | OPA3 | TSPAN12 | INPP5E | TTC21B |
| CYP4V2 | OPN1SW | TTC8 | INVS | TUB |
| DHDDS | PAX6 | TTPA | IQCB1 | TUBGCP6 |
| EFEMP1 | PDE6A | TULP1 | JAG1 | UNC119 |
| ELOVL4 | PDE6B | USH2A | KIF11 | USH1C |
| EYS | PDE6C | ZNF513 | KIF7 | USH1G |
| FAM161A | PDE6G | AHI1 | LAMA1 | VCAN |
| FSCN2 | PDE6H | ALMS1 | LRP5 | VPS13B |
| FZD4 | PITPNM3 | ALDH3A2 | LZTFL1 | WDPCP |
| GDF6 | PLA2G5 | ANTXR1 | MFN2 | WDR19 |
| GNAT1 | PRCD | ARL13B | MKKS | WFS1 |
| GNAT2 | PROM1 | ATXN7 | MTTP | WHRN |
| GNB3 | PRPF3 | B9D1 | MVK | XPNPEP3 |
| GPR179 | PRPF31 | B9D2 | MYO7A | ZNF423 |
| GRK1 | PRPF6 | BBS10 | NBAS | |
| GRM6 | PRPF8 | BBS12 | NEK8 | |

| | | | |
|---|---|---|---|
| GUCA1A | PRPH2 | BBS4 | NPHP1 |
| GUCA1B | RAX2 | BBS5 | NPHP3 |
| | RB1 | BBS7 | NPHP4 |

**Table S9.** Genes classified as involved in other eye related diseases (OERD). The list includes all genes linked with the HPO term “Eye Disease” – HP:0000478, but those included in the Table S8.

| Gene | | | | | | | |
|---|---|---|---|---|---|---|---|
| AAAS | CENPJ | EPHX2 | HSD3B7 | MYH3 | PRKG1 | SPEF2 | ZNF469 |
| AARS2 | CEP152 | EPM2A | HSF4 | MYH8 | PRMT7 | SPG11 | ZNF644 |
| AASS | CEP57 | ERBB3 | HSPD1 | MYH9 | PRNP | SPINT2 | |
| ABCA1 | CEP85L | ERCC1 | HSPG2 | MYLK | PROC | SPR | |
| ABCA12 | CFH | ERCC2 | HTRA1 | MYO18B | PROK2 | SPRED1 | |
| ABCA2 | CFHR1 | ERCC3 | HTT | MYO5A | PROKR2 | SPRY4 | |
| ABCA7 | CFHR3 | ERCC4 | HUWE1 | MYO6 | PROP1 | SPTBN2 | |
| ABCB7 | CFI | ERCC5 | HYDIN | MYO9A | PROS1 | SQSTM1 | |
| ABCC6 | CFL2 | ERCC6 | HYLS1 | MYOC | PRRX1 | SRC | |
| ABCC8 | CHAT | ERCC8 | ICOS | MYOD1 | PRSS12 | SRCAP | |
| ABCC9 | CHD3 | ERLIN2 | IDS | MYPN | PRSS56 | SRD5A3 | |
| ABCD1 | CHD7 | ERMARD | IDUA | MYT1L | PRX | SREBF1 | |
| ABCD4 | CHD8 | ESCO2 | IER3IP1 | NAA10 | PSAP | SRPX2 | |
| ABCG8 | CHEK2 | ESPN | IFIH1 | NAGA | PSAT1 | SRY | |
| ABHD5 | CHMP1A | ESR1 | IFITM5 | NARS2 | PSEN1 | ST14 | |
| ABL1 | CHMP4B | ETFA | IFNG | NCF1 | PSEN2 | ST3GAL3 | |
| ACADS | CHN1 | ETFB | IFT122 | NCF2 | PTCH1 | ST3GAL5 | |
| ACADSB | CHRDL1 | ETFDH | IFT43 | NCF4 | PTCH2 | STAT3 | |
| ACBD5 | CHRNA1 | ETHE1 | IGBP1 | NDE1 | PTF1A | STAT4 | |
| ACO2 | CHRNA3 | EVC | IGF1 | NDN | PTH | STIM1 | |
| ACOX1 | CHRNA7 | EVC2 | IGF1R | NDRG1 | PTH1R | STOX1 | |
| ACSL4 | CHRNB1 | EXOSC3 | IGF2 | NDST1 | PTPN11 | STRA6 | |
| ACTA1 | CHRND | EXOSC9 | IGFBP7 | NDUFA1 | PTPN22 | STRADA | |
| ACTB | CHRNE | EXT2 | IGLL1 | NDUFA10 | PTPN23 | STS | |
| ACTG1 | CHRNG | EXTL3 | IGSF3 | NDUFA11 | PTS | STT3A | |
| ACTN2 | CHST3 | EYA1 | IKZF1 | NDUFA12 | PUF60 | STT3B | |
| ACVR1 | CHST6 | EZH2 | IL10 | NDUFA13 | PUS1 | STX11 | |
| ACY1 | CHSY1 | FA2H | IL11RA | NDUFA2 | PUS3 | STX16 | |
| ADAM17 | CIC | FAM126A | IL12B | NDUFA4 | PWRN1 | STXBP1 | |
| ADAM22 | CITED2 | FAM20C | IL17F | NDUFA6 | PXDN | SUCLA2 | |
| ADAMTS10 | CLCC1 | FAN1 | IL17RA | NDUFA9 | PYCR1 | SUFU | |
| ADAMTS17 | CLCN2 | FANCA | IL1RAPL1 | NDUFAF1 | RAB18 | SUMF1 | |

| | | | | | | |
|---|---|---|---|---|---|---|
| ADAMTS2 | CLCN4 | FANCB | IL23R | NDUFAF2 | RAB23 | SUOX |
| ADAMTSL4 | CLCN7 | FANCC | IL2RA | NDUFAF4 | RAB27A | SUZ12 |
| ADAR | CLCNKB | FANCD2 | IL6 | NDUFAF5 | RAB28 | SYNE1 |
| ADARB1 | CLDN16 | FANCE | IL6ST | NDUFAF6 | RAB39B | SYNE2 |
| ADGRG1 | CLDN19 | FANCF | INPP5K | NDUFB11 | RAB3GAP1 | SYNGAP1 |
| ADK | CLEC7A | FANCG | INPPL1 | NDUFB3 | RAB3GAP2 | SYT14 |
| ADNP | CNBP | FANCI | INS | NDUFB9 | RAC1 | SYT2 |
| ADSL | CNKSR2 | FANCL | INSR | NDUFS1 | RAD21 | TAB2 |
| AGA | CNOT3 | FANCM | IPW | NDUFS2 | RAD50 | TAC3 |
| AGK | CNTN1 | FARS2 | IQSEC2 | NDUFS4 | RAD51 | TACO1 |
| AGL | COA3 | FAS | IRX5 | NDUFS6 | RAD51C | TACR3 |
| Aug-02 | COG1 | FASLG | ISCA2 | NDUFS7 | RAF1 | TACSTD2 |
| AGRN | COG4 | FASTKD2 | ITCH | NDUFV1 | RAG1 | TAF1 |
| AGTPBP1 | COG5 | FBLN1 | ITGA2B | NDUFV2 | RAG2 | TAF2 |
| AGXT | COG6 | FBLN5 | ITGA3 | NEB | RAI1 | TARDBP |
| AHCY | COL12A1 | FBN2 | ITGA7 | NEDD4L | RALGAPA1 | TAT |
| AHSG | COL17A1 | FBXO11 | ITGB3 | NEFL | RANBP2 | TBC1D23 |
| AIFM1 | COL18A1 | FBXO7 | ITGB6 | NEK1 | RAPSN | TBC1D24 |
| AIMP1 | COL1A1 | FBXW11 | ITPA | NF1 | RARS2 | TBCD |
| AIP | COL1A2 | FDFT1 | ITPR1 | NFIX | RASGRP1 | TBCE |
| AIRE | COL25A1 | FERMT1 | JAK2 | NFKB2 | RAX | TBK1 |
| AKT1 | COL4A1 | FGD1 | JAM3 | NGF | RBBP8 | TBL1XR1 |
| AKT3 | COL4A2 | FGF10 | KANK1 | NGLY1 | RBM10 | TBP |
| ALDH18A1 | COL4A3 | FGF14 | KANSL1 | NHLRC1 | RBM8A | TBX1 |
| ALDH5A1 | COL4A4 | FGF20 | KAT5 | NHP2 | RBPJ | TBX15 |
| ALDH6A1 | COL4A5 | FGF3 | KAT6A | NHS | RECQL4 | TBX2 |
| ALDH7A1 | COL4A6 | FGF8 | KAT6B | NIN | RELN | TBX22 |
| ALDOA | COL5A1 | FGF9 | KBTBD13 | NIPAL4 | RFT1 | TBX4 |
| ALDOB | COL5A2 | FGFR1 | KCNA1 | NIPBL | RIC1 | TCF3 |
| ALG1 | COL6A1 | FGFR2 | KCNA4 | NKX2-5 | RIN2 | TCF4 |
| ALG11 | COL6A2 | FGFR3 | KCNC3 | NKX2-6 | RIPK4 | TCIRG1 |
| ALG12 | COL6A3 | FGFRL1 | KCND3 | NKX3-2 | RNASEH2A | TCOF1 |
| ALG13 | COL7A1 | FH | KCNJ1 | NLRP1 | RNASEH2B | TCTN3 |
| ALG14 | COL8A2 | FHL1 | KCNJ11 | NLRP3 | RNASET2 | TDO2 |
| ALG2 | COLEC10 | FIG4 | KCNJ2 | NME8 | RNF168 | TDP1 |
| ALG3 | COLEC11 | FKBP10 | KCNJ6 | NOD2 | ROBO3 | TDRD7 |
| ALG6 | COLQ | FKBP14 | KCNMA1 | NODAL | ROR2 | TECR |
| ALG8 | COMT | FKRP | KCNN3 | NOG | RORA | TEK |
| ALG9 | COQ2 | FKTN | KCNQ3 | NOP56 | RPIA | TELO2 |
| ALOX12B | COQ5 | FLCN | KCNQ5 | NOTCH1 | RPL11 | TERT |
| ALOXE3 | COQ8A | FLI1 | KCTD7 | NOTCH2 | RPL35A | TET2 |

| | | | | | | |
|---|---|---|---|---|---|---|
| ALPL | CORIN | FLNA | KDM5C | NOTCH3 | RPL5 | TFAP2A |
| ALS2 | COX10 | FLNB | KDM6A | NPAP1 | RPS10 | TFAP2B |
| ALX1 | COX14 | FLT4 | KDM6B | NPC1 | RPS19 | TFG |
| ALX3 | COX15 | FMN2 | KERA | NPC2 | RPS24 | TFRC |
| ALX4 | COX6B1 | FMR1 | KIF1A | NPM1 | RPS26 | TGFB1 |
| AMACR | COX7B | FOXC1 | KIF1B | NRAS | RPS28 | TGFB2 |
| AMER1 | CP | FOXC2 | KIF21A | NRXN1 | RPS6KA3 | TGFB3 |
| ANAPC1 | CPT2 | FOXE3 | KIF5A | NSD1 | RPS7 | TGFBI |
| ANK1 | CR2 | FOXG1 | KISS1 | NSD2 | RRAS2 | TGIF1 |
| ANKH | CRADD | FOXJ1 | KISS1R | NSDHL | RRM2B | TGM1 |
| ANKRD11 | CRBN | FOXP1 | KIT | NSMF | RSPH4A | TGM5 |
| ANO10 | CREBBP | FOXRED1 | KITLG | NSUN2 | RSPH9 | TGM6 |
| ANOS1 | CRLF1 | FRAS1 | KLF11 | NTF4 | RSRC1 | TH |
| AP1S1 | CRTAP | FREM1 | KLHL41 | NTRK1 | RTTN | THOC2 |
| AP1S2 | CRYAA | FREM2 | KMT2C | NTRK2 | RUBCN | THPO |
| AP3B1 | CRYAB | FRG1 | KMT2D | NUBPL | RUNX2 | THRA |
| AP3B2 | CRYBA1 | FRMD7 | KMT2E | NUP62 | SACS | THRB |
| AP3D1 | CRYBA4 | FRMPD4 | KNL1 | OCA2 | SALL1 | TK2 |
| AP4B1 | CRYBB1 | FTL | KRAS | OCLN | SALL4 | TMCO1 |
| AP4E1 | CRYBB2 | FUCA1 | KRIT1 | OCRL | SAMD9 | TMEM231 |
| AP4S1 | CRYBB3 | FUS | KRT1 | ODC1 | SAMHD1 | TMEM70 |
| AP5Z1 | CRYGB | FUT8 | KRT10 | OGT | SAR1B | TMTC3 |
| APOA1 | CRYGC | FXN | KRT12 | OPHN1 | SARDH | TNFAIP3 |
| APOC2 | CRYGD | FYCO1 | KRT14 | OPN1LW | SATB2 | TNFRSF11A |
| APOE | CRYGS | GABRA1 | KRT3 | OPN1MW | SBDS | TNFRSF11B |
| APP | CSF1R | GABRA2 | KRT5 | OPTN | SBF2 | TNFRSF13B |
| APTX | CSGALNACT1 | GABRA5 | KRT74 | ORAI1 | SC5D | TNFRSF13C |
| ARHGAP31 | CST3 | GABRB3 | KRT83 | ORC1 | SCARF2 | TNFRSF1A |
| ARHGDIA | CST6 | GABRD | KRT86 | OSMR | SCN1A | TNFSF11 |
| ARID1A | CSTA | GABRG2 | L1CAM | OSTM1 | SCN1B | TNFSF4 |
| ARID1B | CTC1 | GAD1 | L2HGDH | P3H1 | SCN2A | TOR1A |
| ARSA | CTDP1 | GALC | LAMA2 | P3H2 | SCN3A | TP53RK |
| ARSB | CTLA4 | GALK1 | LAMA3 | PACS1 | SCN4A | TP63 |
| ARX | CTNNB1 | GALNS | LAMB1 | PAFAH1B1 | SCN8A | TPI1 |
| ASAH1 | CTNND1 | GALNT2 | LAMB2 | PAH | SCN9A | TPM2 |
| ASB10 | CTNS | GALNT3 | LAMB3 | PARK7 | SCP2 | TPM3 |
| ASNS | CTSA | GALT | LAMC2 | PAX1 | SCYL1 | TRAF3IP1 |
| ASPA | CTSK | GAN | LAMC3 | PAX3 | SDHA | TRAF3IP2 |
| ASPM | CYB5A | GAS2L2 | LAMP2 | PAX4 | SDHAF1 | TRAK1 |
| ASXL1 | CYB5R3 | GATA1 | LARGE1 | PAX7 | SEC23A | TRAPPC2 |
| ATCAY | CYBA | GATA2 | LARS2 | PBX1 | SEC23B | TRAPPC9 |

| | | | | | | |
|---|---|---|---|---|---|---|
| ATIC | CYBB | GATA3 | LAS1L | PCK1 | SELENON | TREM2 |
| ATM | CYP1B1 | GATA4 | LBR | PCLO | SEMA3A | TRIM37 |
| ATN1 | CYP24A1 | GATA5 | LCAT | PCNT | SEMA3E | TRIO |
| ATOH7 | CYP27A1 | GATA6 | LDLRAP1 | PDCD1 | SEPSECS | TRIP11 |
| ATP13A2 | CYP7B1 | GBA | LEMD3 | PDCD10 | SERAC1 | TRIP12 |
| ATP1A2 | DAG1 | GCDH | LETM1 | PDE4D | SERPINC1 | TRMT1 |
| ATP1A3 | DARS2 | GCH1 | LGI4 | PDGFB | SERPING1 | TRPV3 |
| ATP2B2 | DBH | GCK | LHX3 | PDGFRB | SERPINH1 | TRPV4 |
| ATP2B3 | DCC | GCNT2 | LHX4 | PDHA1 | SERPINI1 | TRRAP |
| ATP6AP1 | DCN | GDF3 | LIFR | PDHB | SETBP1 | TSEN2 |
| ATP6V0A2 | DCTN1 | GDF5 | LIG4 | PDHX | SETD2 | TSEN34 |
| ATP8A2 | DCX | GDNF | LIM2 | PDP1 | SETX | TSFM |
| ATR | DDB2 | GEMIN4 | LINS1 | PDSS1 | SF3B1 | TSHR |
| ATRX | DDC | GFM1 | LIPH | PDSS2 | SF3B4 | TSR2 |
| ATXN1 | DDHD2 | GFPT1 | LMBR1 | PDX1 | SH2B3 | TTBK2 |
| ATXN10 | DDOST | GGCX | LMNB1 | PDXK | SH3BP2 | TTC19 |
| ATXN2 | DDR2 | GHR | LMOD3 | PDYN | SH3PXD2B | TTC37 |
| ATXN3 | DDX11 | GIGYF2 | LMX1B | PEPD | SH3TC2 | TTI2 |
| ATXN8OS | DDX3X | GJA1 | LOX | PEX10 | SHANK3 | TTR |
| AUH | DDX58 | GJA3 | LOXL1 | PEX11B | SHH | TUBB2B |
| AUTS2 | DEAF1 | GJA5 | LPL | PEX12 | SHOC2 | TUBB3 |
| AVP | DGCR2 | GJA8 | LRBA | PEX13 | SHROOM4 | TUBB6 |
| B3GALNT2 | DGCR6 | GJB1 | LRP1 | PEX14 | SIGMAR1 | TUBGCP4 |
| B3GLCT | DGCR8 | GJB2 | LRP2 | PEX16 | SIK3 | TUSC3 |
| B4GALNT1 | DGUOK | GJB3 | LRP4 | PEX19 | SIL1 | TWIST1 |
| B4GALT7 | DHCR24 | GJB6 | LRPAP1 | PEX26 | SIX1 | TWIST2 |
| BANF1 | DHCR7 | GJC2 | LRPPRC | PEX3 | SIX3 | TWNK |
| BAP1 | DHODH | GK | LRRC8A | PEX5 | SIX6 | TXN2 |
| BCAP31 | DHX37 | GLB1 | LRRK2 | PGAP2 | SKI | TYMP |
| BCL10 | DIAPH1 | GLE1 | LTBP2 | PGAP3 | SLC12A3 | TYR |
| BCL11A | DICER1 | GLI1 | LTBP4 | PHF21A | SLC12A6 | TYRP1 |
| BCOR | DIS3L2 | GLI2 | LYST | PHF6 | SLC16A12 | UBA1 |
| BCORL1 | DKC1 | GLI3 | MADD | PHGDH | SLC16A2 | UBA5 |
| BCR | DLAT | GLIS3 | MAF | PHIP | SLC17A5 | UBE2A |
| BCS1L | DLD | GLRB | MAFA | PHOX2A | SLC19A2 | UBE3A |
| BEAN1 | DLG3 | GLRX5 | MAFB | PHOX2B | SLC19A3 | UBIAD1 |
| BFSP1 | DLG4 | GM2A | MAGEL2 | PI4KA | SLC1A2 | UBR1 |
| BFSP2 | DLL1 | GNAI3 | MALT1 | PIEZO2 | SLC1A3 | UCHL1 |
| BGN | DLX5 | GNAQ | MAN1B1 | PIGA | SLC20A2 | UGP2 |
| BIN1 | DMPK | GNAS | MAN2B1 | PIGL | SLC24A5 | UGT1A1 |
| BLK | DNAAF1 | GNB5 | MANBA | PIGN | SLC25A1 | UMPS |

| | | | | | | |
|---|---|---|---|---|---|---|
| BLM | DNAAF2 | GNE | MAP2K1 | PIGO | SLC25A13 | UNC80 |
| BLNK | DNAAF3 | GNPAT | MAP2K2 | PIGV | SLC25A15 | UQCRFS1 |
| BLOC1S5 | DNAAF5 | GNPTAB | MAPK1 | PIK3CA | SLC25A19 | UROC1 |
| BLOC1S6 | DNAH11 | GNRH1 | MAPKAPK3 | PIK3CD | SLC25A20 | UROS |
| BMP1 | DNAH5 | GNRHR | MAPT | PIK3R1 | SLC25A22 | USB1 |
| BMP2 | DNAH9 | GORAB | MARS2 | PIK3R2 | SLC25A4 | USP7 |
| BMP4 | DNAI1 | GP1BA | MASP1 | PIK3R5 | SLC26A2 | USP9X |
| BMPER | DNAI2 | GP1BB | MBD5 | PIKFYVE | SLC29A3 | VANGL2 |
| BNC2 | DNAJC19 | GPC3 | MBOAT7 | PINK1 | SLC2A1 | VAX1 |
| BOLA3 | DNAJC6 | GPC4 | MBTPS2 | PITX1 | SLC2A10 | VCP |
| BRAF | DNAL1 | GPIHBP1 | MCM3AP | PITX2 | SLC33A1 | VIM |
| BRAT1 | DNASE1L3 | GPR143 | MCM5 | PITX3 | SLC35A1 | VLDLR |
| BRIP1 | DNM1 | GRHL2 | MCOLN1 | PLA2G6 | SLC35A2 | VMA21 |
| BRPF1 | DNM1L | GRIA3 | MECP2 | PLCB4 | SLC35C1 | VPS13A |
| BTK | DNM2 | GRID2 | MED12 | PLCD1 | SLC36A2 | VPS35 |
| BUB1 | DNMT1 | GRIK2 | MED13 | PLCG2 | SLC37A4 | VSX1 |
| BUB1B | DNMT3A | GRIN1 | MED13L | PLEC | SLC39A13 | VSX2 |
| C19orf12 | DNMT3B | GRIN2A | MED23 | PLEKHG2 | SLC39A4 | WAS |
| C1QTNF5 | DOCK3 | GRIN2B | MED25 | PLEKHM1 | SLC3A1 | WASHC4 |
| C1R | DOCK6 | GRIP1 | MEF2C | PLG | SLC40A1 | WASHC5 |
| C9orf72 | DOCK7 | GRM1 | MEFV | PLOD1 | SLC45A2 | WDFY3 |
| CA2 | DOCK8 | GRM7 | MEGF8 | PLOD3 | SLC4A11 | WDR11 |
| CA8 | DOK7 | GSC | MEIS2 | PLP1 | SLC4A4 | WDR35 |
| CACNA1A | DPAGT1 | GSN | METTL5 | PMM2 | SLC52A2 | WDR36 |
| CACNA1D | DPM1 | GSR | MFF | PMP22 | SLC52A3 | WDR4 |
| CACNA1E | DPP6 | GSS | MGP | PMS1 | SLC5A7 | WDR45 |
| CACNA1G | DPYD | GTF2H5 | MID1 | PNKD | SLC6A19 | WDR45B |
| CACNA1H | DSE | GUSB | MIF | PNPLA1 | SLC6A20 | WDR81 |
| CACNA2D2 | DSG4 | HACE1 | MIP | PNPO | SLC6A3 | WIPF1 |
| CACNB4 | DST | HADH | MIR184 | PNPT1 | SLC6A8 | WNK1 |
| CACNG2 | DTNBP1 | HADHA | MITF | POC1A | SLC6A9 | WNT10A |
| CALR | DVL1 | HADHB | MKRN3 | POGZ | SLCO2A1 | WNT10B |
| CAMK2G | DYNC1H1 | HBA2 | MLH3 | POLD1 | SLITRK6 | WNT3 |
| CAMTA1 | DYNC2LI1 | HBB | MLPH | POLG | SLX4 | WNT5A |
| CANT1 | DYRK1A | HCCS | MLXIPL | POLG2 | SMARCA4 | WRAP53 |
| CASK | EARS2 | HCN1 | MMADHC | POLH | SMARCAL1 | WRN |
| CASP10 | EBP | HCRT | MME | POLR1C | SMARCB1 | WWOX |
| CASR | EDN1 | HDAC4 | MMP1 | POLR1D | SMARCE1 | XPA |
| CAV1 | EDN3 | HDAC6 | MMP14 | POLR3A | SMC1A | XPC |
| CBL | EDNRA | HDAC8 | MMP2 | POLR3B | SMC3 | XRCC1 |
| CBS | EDNRB | HERC2 | MN1 | POMGNT2 | SMCHD1 | XRCC2 |

| | | | | | | |
|---|---|---|---|---|---|---|
| CC2D1A | EFEMP2 | HESX1 | MOCS1 | POMK | SMO | XRCC4 |
| CCBE1 | EFHC1 | HEXA | MOCS2 | POMT1 | SMOC1 | XYLT1 |
| CCDC22 | EFNB1 | HEXB | MOGS | POMT2 | SMPD1 | XYLT2 |
| CCDC39 | EGFR | HGD | MPC1 | POR | SMS | YAP1 |
| CCDC40 | EGR2 | HIBCH | MPDZ | PORCN | SNAI2 | YARS2 |
| CCDC88C | EHMT1 | HLCS | MPL | POU1F1 | SNAP25 | YIF1B |
| CCM2 | EIF2AK3 | HMGA2 | MPV17 | POU3F4 | SNAP29 | YY1 |
| CCND1 | EIF2B1 | HNF4A | MPZ | POU6F2 | SNCA | ZBTB16 |
| CD19 | EIF2B2 | HNMT | MRE11 | PPIB | SNIP1 | ZBTB18 |
| CD247 | EIF2B3 | HNRNPU | MSMO1 | PPM1D | SNORD115-1 | ZBTB24 |
| CD27 | EIF2B4 | HOXA1 | MSX2 | PPP1R17 | SNORD116-1 | ZC3H14 |
| CD79A | EIF2B5 | HOXA13 | MTAP | PPP2R1A | SNRPN | ZDHHC9 |
| CD79B | EIF2S3 | HOXB1 | MTFMT | PPP2R2B | SNX10 | ZEB1 |
| CD96 | EIF4G1 | HPGD | MTHFR | PPP3CA | SOBP | ZEB2 |
| CDC42 | ELN | HPS1 | MTHFS | PQBP1 | SOS1 | ZFHX4 |
| CDH1 | ELP4 | HPS3 | MTM1 | PRDM16 | SOST | ZFPM2 |
| CDH15 | EMD | HPS4 | MTMR14 | PRDM5 | SOX2 | ZFYVE26 |
| CDK4 | ENPP1 | HPS5 | MTO1 | PRDX1 | SOX3 | ZIC1 |
| CDK5RAP2 | ENTPD1 | HPS6 | MTPAP | PREPL | SOX5 | ZIC2 |
| CDKL5 | EP300 | HRAS | MTR | PRF1 | SOX6 | ZIC3 |
| CDKN1C | EPAS1 | HS2ST1 | MTRR | PRICKLE3 | SOX9 | ZMPSTE24 |
| CDKN2A | EPB41L1 | HS6ST1 | MUSK | PRKAR1A | SPAST | ZMYND11 |
| CDON | EPCAM | HSD11B2 | MYF5 | PRKCG | SPATA5 | ZNF335 |
| CEL | EPHA2 | HSD17B4 | MYH2 | PRKDC | SPECC1L | ZNF365 |

**Table S10**. Genes in the database of allelic frequencies not involved in an eye related disease (NRD).

| Gene | | | | | | |
|---|---|---|---|---|---|---|
| A2M | CCRL2 | EPOR | IDH2 | MSH3 | PRMT3 | SRGAP2 |
| A4GALT | CCT5 | EPX | IDO1 | MSH4 | PRMT9 | SRGAP3 |
| A4GNT | CD109 | ERAP1 | IFI30 | MSMB | PRND | SRI |
| AADAC | CD14 | ERAP2 | IFI44L | MSR1 | PRODH | SRP72 |
| AADACL2 | CD151 | ERBB2 | IFITM3 | MSRA | PROK1 | SRPX |
| AAGAB | CD177 | ERBB4 | IFNA10 | MSRB3 | PROKR1 | SRR |
| AARS | CD1A | ERI2 | IFNA17 | MST1R | PROX1 | SSH1 |
| ABAT | CD1E | ERMAP | IFNA2 | MSTN | PROZ | SSPN |
| ABCA10 | CD200 | ERRFI1 | IFNAR1 | MSX1 | PRPH | SST |
| ABCA13 | CD207 | ESAM | IFNAR2 | MT1A | PRSS1 | SSTR5 |
| ABCA3 | CD209 | ESR2 | IFNGR1 | MT2A | PRSS8 | SSX7 |

| | | | | | | |
|---|---|---|---|---|---|---|
| ABCB1 | CD22 | ESRRB | IFNGR2 | MTA1 | PRTG | ST3GAL1 |
| ABCB11 | CD226 | ESRRG | IFNL3 | MTA2 | PSCA | ST3GAL2 |
| ABCB4 | CD244 | ETNPPL | IFRD1 | MTCH2 | PSMA6 | ST3GAL4 |
| ABCC1 | CD2AP | ETS1 | IFT88 | MTHFD1 | PSMC2 | ST3GAL6 |
| ABCC11 | CD320 | ETV4 | IGF2BP2 | MTHFD1L | PSMC3IP | ST5 |
| ABCC2 | CD36 | ETV6 | IGF2R | MTMR2 | PSMD7 | ST6GAL1 |
| ABCC3 | CD38 | EVI5 | IGFALS | MTMR9 | PSPH | ST6GAL2 |
| ABCC4 | CD3D | EWSR1 | IGFBP1 | MTNR1A | PSTPIP1 | ST6GALNAC1 |
| ABCD3 | CD3E | EXO1 | IGFBP3 | MTNR1B | PSTPIP2 | ST6GALNAC2 |
| ABCG1 | CD3G | EXO5 | IGFBP5 | MTSS1 | PTAFR | ST6GALNAC3 |
| ABCG2 | CD4 | EXOC4 | IGHMBP2 | MTUS1 | PTCHD1 | ST6GALNAC4 |
| ABI3BP | CD40 | EXPH5 | IGSF1 | MUC1 | PTCHD3 | ST6GALNAC5 |
| ABL2 | CD40LG | EXT1 | IHH | MUC13 | PTCSC3 | ST6GALNAC6 |
| ABO | CD44 | EXTL1 | IKBIP | MUC15 | PTGDR | ST7 |
| ACACA | CD46 | EXTL2 | IKBKAP | MUC2 | PTGDR2 | ST8SIA1 |
| ACACB | CD5 | EYA4 | IKBKB | MUC3A | PTGDS | ST8SIA2 |
| ACAD10 | CD55 | F10 | IKZF3 | MUC4 | PTGER2 | ST8SIA3 |
| ACAD11 | CD58 | F11 | IL10RA | MUC5B | PTGER4 | ST8SIA4 |
| ACAD8 | CD59 | F12 | IL10RB | MUC6 | PTGES2 | ST8SIA5 |
| ACAD9 | CD72 | F13A1 | IL11 | MUC7 | PTGIR | ST8SIA6 |
| ACADL | CD74 | F13B | IL12A | MURC | PTGIS | STAR |
| ACADM | CD80 | F2 | IL12RB1 | MUS81 | PTGS1 | STARD9 |
| ACADVL | CD81 | F2R | IL12RB2 | MUT | PTGS2 | STAT1 |
| ACAN | CD86 | F2RL1 | IL13 | MX1 | PTHLH | STAT5B |
| ACAT1 | CD8A | F3 | IL16 | MXI1 | PTK7 | STAT6 |
| ACAT2 | CDA | F5 | IL17A | MYB | PTPN1 | STEAP3 |
| ACBD6 | CDAN1 | F7 | IL17RB | MYBL2 | PTPN12 | STEAP4 |
| ACCS | CDC42BPB | F8 | IL17REL | MYBPC1 | PTPN13 | STH |
| ACE | CDC6 | F9 | IL18 | MYC | PTPN14 | STIL |
| ACHE | CDC73 | FAAH | IL18R1 | MYCL | PTPN2 | STK10 |
| ACKR1 | CDCA7L | FAAH2 | IL18RAP | MYCN | PTPN21 | STK11IP |
| ACLY | CDH12 | FABP1 | IL19 | MYEF2 | PTPN6 | STK3 |
| ACP1 | CDH13 | FABP2 | IL1A | MYF6 | PTPRB | STK32A |
| ACP5 | CDH5 | FABP3 | IL1B | MYH13 | PTPRC | STK33 |
| ACSF3 | CDH8 | FABP4 | IL1R1 | MYH14 | PTPRD | STK35 |
| ACSL5 | CDK11A | FABP6 | IL1RL1 | MYH15 | PTPRF | STK36 |
| ACSL6 | CDK16 | FABP7 | IL1RN | MYH6 | PTPRJ | STK39 |
| ACSM2B | CDK5R1 | FADD | IL2 | MYL1 | PTPRK | STK4 |

| | | | | | | |
|---|---|---|---|---|---|---|
| ACSM3 | CDK5RAP3 | FADS2 | IL20RA | MYLIP | PTPRN2 | STMN1 |
| ACTN3 | CDK6 | FAH | IL20RB | MYLK2 | PTPRO | STRC |
| ACTN4 | CDK7 | FAM104A | IL21 | MYO15A | PTPRQ | STX1A |
| ACVR1B | CDKAL1 | FAM120A | IL21R | MYO1A | PTPRT | STXBP2 |
| ACVR1C | CDKL3 | FAM134B | IL2RG | MYO1C | PTRF | STXBP5 |
| ACVR2A | CDKN1A | FAM161B | IL3 | MYO1E | PUS10 | SUCLG1 |
| ACVR2B | CDKN1B | FAM205A | IL31RA | MYO1F | PVR | SUCO |
| ADA | CDKN2B | FAM20A | IL36RN | MYO3A | PVT1 | SUGCT |
| ADAM10 | CDKN2B-AS1 | FAM47B | IL4 | MYO5B | PYCRL | SULF1 |
| ADAM12 | CDKN2C | FAM58A | IL4R | MYO5C | PYGB | SULT1A1 |
| ADAM19 | CDT1 | FAM83H | IL5 | MYO7B | PYGL | SULT1C2 |
| ADAM23 | CDX2 | FAM8A1 | IL6R | MYO9B | PYGM | SULT1E1 |
| ADAM33 | CEACAM16 | FAM91A1 | IL7 | MYOCD | PYY | SULT2A1 |
| ADAM7 | CEBPA | FASN | IL7R | MYOM1 | PZP | SULT2B1 |
| ADAMTS1 | CEBPE | FBLIM1 | IL9 | MYOT | QDPR | SULT4A1 |
| ADAMTS13 | CECR2 | FBN3 | ILDR1 | MYOZ2 | QKI | SUMO1 |
| ADAMTS16 | CELSR1 | FBP1 | ILK | MYT1 | RAB11FIP5 | SUMO4 |
| ADAMTSL2 | CELSR2 | FBXO10 | IMMP2L | NAGLU | RAB25 | SUN2 |
| ADAMTSL3 | CEMIP | FBXO18 | IMMT | NAGPA | RAB27B | SUPT16H |
| ADCY10 | CENPO | FBXW4 | IMPA2 | NAIP | RAB29 | SV2B |
| ADCY3 | CENPP | FBXW7 | IMPAD1 | NAMPT | RAB2A | SYCE2 |
| ADCY5 | CEP135 | FCAR | IMPDH2 | NAT1 | RAB40AL | SYCP3 |
| ADCY6 | CEP63 | FCER1A | INF2 | NAT2 | RAB7A | SYK |
| ADCY9 | CEP68 | FCER2 | ING1 | NAT8L | RABGGTA | SYN1 |
| ADCYAP1 | CER1 | FCGR1A | INMT | NAV2 | RABL6 | SYN2 |
| ADD1 | CERS6 | FCGR2A | INPP4A | NBEA | RAC2 | SYN3 |
| ADD2 | CES1 | FCGR2B | INPP5B | NBEAL2 | RAD21L1 | SYNGR1 |
| ADH1B | CES2 | FCGR3A | INPP5D | NBN | RAD23B | SYNM |
| ADH1C | CETP | FCGR3B | INSIG1 | NBPF1 | RAD51B | SYNPO |
| ADH4 | CFAP53 | FCGRT | INSIG2 | NCALD | RAD51D | SYP |
| ADH5 | CFAP57 | FCN2 | INSL3 | NCAM1 | RAD52 | SYT11 |
| ADH7 | CFC1 | FCN3 | INSL6 | NCAN | RAD54B | SYTL3 |
| ADIPOQ | CFD | FCRL3 | IQGAP1 | NCAPD2 | RAD54L | SYTL5 |
| ADM | CFHR2 | FECH | IQGAP2 | NCKAP1 | RAD9A | T |
| ADORA1 | CFHR4 | FEM1A | IQGAP3 | NCOA1 | RAET1L | TAAR1 |
| ADORA2A | CFHR5 | FEM1B | IRAK1 | NCOA3 | RALGDS | TAAR6 |
| ADORA3 | CFLAR | FEN1 | IRAK3 | NCOA4 | RANGRF | TAAR9 |
| ADRA1A | CFP | FERMT3 | IRAK4 | NCS1 | RAP1GDS1 | TAF15 |

| | | | | | | |
|---|---|---|---|---|---|---|
| ADRA2A | CFTR | FEV | IRF1 | NCSTN | RARA | TAF1C |
| ADRA2B | CGA | FEZF2 | IRF2 | NDOR1 | RASA1 | TAF1L |
| ADRA2C | CGB3 | FFAR1 | IRF4 | NDST2 | RASGRP2 | TAF7L |
| ADRB1 | CHD1L | FFAR4 | IRF5 | NDST3 | RASSF1 | TAL1 |
| ADRB2 | CHD2 | FGA | IRF6 | NDST4 | RASSF5 | TAL2 |
| ADRB3 | CHD6 | FGB | IRF8 | NDUFA7 | RB1CC1 | TALDO1 |
| ADTRP | CHDH | FGD3 | IRGM | NDUFA8 | RBFOX1 | TAS1R1 |
| AFF2 | CHFR | FGD4 | IRS1 | NDUFAF7 | RBL1 | TAS1R2 |
| AFF3 | CHGA | FGF1 | IRS2 | NDUFB1 | RBL2 | TAS2R16 |
| AFP | CHGB | FGF2 | IRS4 | NDUFB6 | RBM15 | TAS2R3 |
| AGBL4 | CHI3L1 | FGF23 | IRX4 | NDUFS5 | RBM20 | TAS2R38 |
| AGGF1 | CHI3L2 | FGFBP1 | ISCU | NDUFV3 | RBM28 | TAS2R9 |
| AGMO | CHIA | FGFR4 | ISL1 | NEBL | RBMXL2 | TAZ |
| AGO1 | CHIC2 | FGG | ISPD | NECTIN1 | RC3H1 | TBC1D1 |
| AGPAT2 | CHIT1 | FHIT | ITGA1 | NECTIN4 | RCAN1 | TBC1D4 |
| AGPS | CHL1 | FHL2 | ITGA11 | NEDD4 | RDH8 | TBC1D9 |
| AGRP | CHMP2B | FIGLA | ITGA2 | NEDD9 | RDX | TBL1X |
| AGT | CHPF2 | FIP1L1 | ITGA4 | NEFH | REEP1 | TBL1Y |
| AGTR1 | CHRD | FKBP1A | ITGA6 | NEFM | REL | TBX10 |
| AGTR2 | CHRFAM7A | FKBP4 | ITGA9 | NEGR1 | REN | TBX19 |
| AGXT2 | CHRM1 | FKBP5 | ITGAE | NEIL1 | REPS2 | TBX20 |
| AHRR | CHRM2 | FKBP6 | ITGAM | NEIL2 | RETN | TBX21 |
| AHSP | CHRM3 | FKBP8 | ITGB1 | NELFA | REV3L | TBX3 |
| AICDA | CHRNA2 | FLG | ITGB2 | NELL1 | RFC2 | TBX5 |
| AK1 | CHRNA4 | FLT1 | ITGB4 | NEU2 | RFWD2 | TBXA2R |
| AK2 | CHRNA5 | FLT3 | ITIH1 | NEUROG3 | RFX2 | TBXAS1 |
| AK7 | CHRNA9 | FLVCR2 | ITIH3 | NEXN | RFX5 | TCF21 |
| AK8 | CHRNB2 | FMN1 | ITIH4 | NFATC2 | RFX6 | TCF7 |
| AKAP10 | CHRNB4 | FMO1 | ITIH6 | NFATC3 | RFX8 | TCF7L1 |
| AKAP13 | CHST7 | FMO2 | ITK | NFATC4 | RFXANK | TCF7L2 |
| AKAP9 | CHST8 | FMO3 | ITPKC | NFE2L1 | RFXAP | TCN1 |
| AKR1B1 | CHSY3 | FMO4 | ITPR3 | NFE2L2 | RGMA | TCN2 |
| AKR1C2 | CHUK | FMO5 | ITSN2 | NFIA | RGS2 | TCP1 |
| AKR1C3 | CIAPIN1 | FMO6P | IVD | NFKB1 | RGS6 | TCTE1 |
| AKR1C4 | CIDEA | FMOD | IYD | NFKBIA | RGS7 | TCTE3 |
| AKR1D1 | CIDEC | FN1 | JAG2 | NFKBIZ | RHAG | TDGF1 |
| AKR7A2 | CIITA | FN3K | JAK3 | NFU1 | RHBDF2 | TEC |
| AKR7A3 | CILP | FOLH1 | JMJD1C | NGFR | RHCE | TECPR2 |

| | | | | | | |
|---|---|---|---|---|---|---|
| AKT2 | CKM | FOLR1 | JPH2 | NHEJ1 | RHD | TECTA |
| ALAD | CLCA1 | FOXA2 | JPH3 | NICN1 | RHOB | TEKT2 |
| ALAS2 | CLCA2 | FOXA3 | JRK | NID1 | RHOH | TENM4 |
| ALB | CLCF1 | FOXD3 | JUN | NINJ1 | RHPN2 | TEP1 |
| ALCAM | CLCN1 | FOXE1 | JUNB | NIP7 | RIC3 | TET1 |
| ALDH16A1 | CLCN3 | FOXF1 | JUP | NIPA1 | RIMS3 | TEX14 |
| ALDH1A1 | CLCN5 | FOXF2 | KALRN | NIPSNAP1 | RIOK2 | TF |
| ALDH1A2 | CLCN6 | FOXI1 | KARS | NIPSNAP3A | RMI1 | TFAM |
| ALDH2 | CLCNKA | FOXK1 | KAT2A | NKAIN2 | RMND1 | TFB1M |
| ALDH4A1 | CLDN1 | FOXM1 | KATNAL2 | NKX2-1 | RNASE3 | TFCP2 |
| ALG10B | CLDN14 | FOXN1 | KCNA3 | NKX2-3 | RNASEL | TFE3 |
| ALG5 | CLEC11A | FOXO1 | KCNA5 | NKX3-1 | RNF114 | TFF1 |
| ALK | CLEC2D | FOXP2 | KCNA6 | NLGN1 | RNF135 | TFPI |
| ALOX12 | CLEC3B | FOXP3 | KCNAB1 | NLGN2 | RNF139 | TFR2 |
| ALOX15 | CLEC4M | FPGS | KCNAB2 | NLGN3 | RNF170 | TG |
| ALOX5 | CLIC2 | FPR1 | KCND2 | NLGN4X | RNF212 | TGFBR3 |
| ALOX5AP | CLIP2 | FPR2 | KCNE1 | NLGN4Y | RNF213 | TGFBRAP1 |
| ALS2CL | CLK2 | FRA10AC1 | KCNE2 | NLRP12 | RNF6 | TGM2 |
| AMBN | CLMP | FREM3 | KCNE3 | NLRP14 | RNLS | THADA |
| AMELX | CLNK | FRK | KCNE4 | NLRP2 | ROBO1 | THAP1 |
| AMELY | CLOCK | FRMD6 | KCNE5 | NLRP7 | ROBO2 | THBD |
| AMH | CLPS | FRY | KCNH3 | NLRX1 | ROCK1 | THBS1 |
| AMHR2 | CLPTM1 | FRZB | KCNIP1 | NMB | ROCK2 | THBS2 |
| AMN | CLPTM1L | FSCB | KCNIP4 | NME1 | ROPN1L | THBS4 |
| AMPD1 | CLSTN2 | FSHB | KCNJ10 | NME5 | ROS1 | THSD7A |
| AMPD3 | CLTCL1 | FSHR | KCNJ12 | NME7 | RPA1 | TICAM1 |
| AMT | CLU | FST | KCNJ15 | NMT2 | RPH3AL | TIMM44 |
| ANG | CLUL1 | FTCD | KCNJ3 | NMU | RPL21 | TIMP1 |
| ANGPT1 | CLYBL | FTHL17 | KCNJ5 | NNT | RPL24 | TIMP2 |
| ANGPT2 | CMA1 | FTO | KCNJ8 | NOBOX | RPL38 | TINAG |
| ANGPTL4 | CMPK1 | FTSJ1 | KCNJ9 | NOD1 | RPN2 | TIRAP |
| ANGPTL5 | CNDP1 | FURIN | KCNK18 | NOP16 | RPS15 | TJP2 |
| ANK2 | CNKSR1 | FUT1 | KCNK3 | NOS1 | RPS3 | TLDC2 |
| ANK3 | CNNM2 | FUT2 | KCNK6 | NOS1AP | RPS6KB1 | TLK1 |
| ANKK1 | CNOT4 | FUT3 | KCNK9 | NOS2 | RPS6KL1 | TLL1 |
| ANKRD1 | CNPY3 | FUT6 | KCNMB3 | NOS3 | RPTOR | TLR1 |
| ANKRD26 | CNR1 | FUZ | KCNMB4 | NPAS2 | RRH | TLR10 |
| ANKS1A | CNR2 | FXYD6 | KCNN2 | NPAS3 | RRM1 | TLR2 |

| | | | | | | |
|---|---|---|---|---|---|---|
| ANKS1B | CNTF | FZD1 | KCNQ2 | NPAT | RRP1B | TLR3 |
| ANKS6 | CNTN4 | FZD3 | KCNQ4 | NPC1L1 | RSC1A1 | TLR4 |
| ANO3 | CNTNAP2 | FZD6 | KCNS1 | NPFFR2 | RSPO1 | TLR5 |
| ANO5 | CNTNAP4 | FZD9 | KCNS3 | NPHS1 | RSPO4 | TLR6 |
| ANO6 | CNTNAP5 | G6PC | KCNT1 | NPHS2 | RTN2 | TLR7 |
| ANO7 | COA5 | G6PC2 | KCNV1 | NPL | RTN4R | TLR8 |
| ANTXR2 | COCH | G6PC3 | KCTD13 | NPPB | RUNX1 | TLR9 |
| ANXA11 | COG2 | G6PD | KDM3A | NPPC | RUNX3 | TLX1 |
| ANXA5 | COG3 | GAB2 | KDM4C | NPR1 | RUVBL1 | TLX2 |
| AOAH | COG7 | GABRA6 | KDM5A | NPR2 | RXFP2 | TLX3 |
| AOC1 | COL10A1 | GABRG1 | KDR | NPR3 | RXRA | TM4SF19 |
| APAF1 | COL6A4P2 | GABRG3 | KEL | NPSR1 | RXRG | TMC1 |
| APBA2 | COL6A5 | GABRR2 | KHDC3L | NPTN | RYK | TMC6 |
| APBB1 | COMMD1 | GAD2 | KHK | NPY | RYR3 | TMC8 |
| APBB2 | COMP | GADD45A | KIAA0100 | NPY1R | S100B | TMEM114 |
| APBB3 | COQ4 | GADD45B | KIAA0232 | NPY2R | S1PR1 | TMEM135 |
| APCDD1 | COQ9 | GAK | KIAA0319 | NQO1 | SAA1 | TMEM165 |
| APH1A | CORO1A | GAL3ST1 | KIAA0513 | NQO2 | SAA2 | TMEM173 |
| APH1B | COX4I1 | GAL3ST2 | KIAA1257 | NR0B1 | SAGE1 | TMEM185A |
| APLNR | COX4I2 | GAL3ST3 | KIAA1462 | NR0B2 | SARS2 | TMEM187 |
| APOA4 | COX7A2 | GAL3ST4 | KIAA2022 | NR1H2 | SART1 | TMEM2 |
| APOA5 | CPA4 | GALNT11 | KIF17 | NR1H3 | SART3 | TMEM249 |
| APOBEC1 | CPA6 | GALNT12 | KIF18A | NR1H4 | SAT1 | TMEM39A |
| APOBEC3B | CPB2 | GALNT13 | KIF1BP | NR1I2 | SATL1 | TMEM52B |
| APOBEC3G | CPE | GALNT14 | KIF22 | NR1I3 | SBNO1 | TMEM8A |
| APOBEC3H | CPLX2 | GALNT18 | KIF27 | NR2E1 | SCAP | TMEM9 |
| APOC1 | CPN1 | GALNT5 | KIF5B | NR2F2 | SCARB1 | TMEM99 |
| APOC3 | CPOX | GALNT6 | KIF6 | NR3C1 | SCARB2 | TMIE |
| APOD | CPS1 | GALNT7 | KIFAP3 | NR3C2 | SCG2 | TMLHE |
| APOH | CPT1A | GALNT8 | KIR2DL1 | NR4A1 | SCG3 | TMPO |
| APOL1 | CPT1B | GALNT9 | KIR2DL3 | NR4A2 | SCGB1A1 | TMPRSS11A |
| APOL3 | CPZ | GALNTL5 | KIR2DL4 | NR4A3 | SCGB1D2 | TMPRSS15 |
| APRT | CR1 | GALNTL6 | KIR3DL1 | NR5A1 | SCGB3A2 | TMPRSS3 |
| AQP1 | CREB1 | GALP | KIR3DL2 | NRCAM | SCLT1 | TMPRSS4 |
| AQP2 | CREB3L3 | GAMT | KIRREL3 | NRG1 | SCN10A | TMPRSS5 |
| AQP3 | CRELD1 | GAP43 | KL | NRG3 | SCN11A | TMPRSS6 |
| AQP4 | CRH | GARS | KLB | NRIP1 | SCN2B | TNC |
| AQP5 | CRHR1 | GAS1 | KLC1 | NRP2 | SCN3B | TNFAIP2 |

| | | | | | | |
|---|---|---|---|---|---|---|
| AQP7 | CRISP2 | GAS6 | KLF10 | NRTN | SCN4B | TNFRSF10A |
| AR | CRK | GATAD1 | KLF5 | NRXN2 | SCN7A | TNFRSF10B |
| AREL1 | CRKL | GATM | KLF6 | NRXN3 | SCNN1A | TNFRSF1B |
| ARFGEF2 | CRP | GBA3 | KLF7 | NSUN7 | SCNN1B | TNFRSF25 |
| ARG1 | CRYM | GBE1 | KLF8 | NT5C1B | SCNN1G | TNFRSF4 |
| ARHGAP24 | CSDE1 | GBGT1 | KLHDC8B | NT5C3A | SCO1 | TNFRSF9 |
| ARHGAP26 | CSF1 | GC | KLHL10 | NT5E | SCRIB | TNFSF10 |
| ARHGAP45 | CSF2 | GCGR | KLHL3 | NTF3 | SCUBE2 | TNFSF13B |
| ARHGAP6 | CSF2RB | GCKR | KLHL9 | NTNG1 | SDC3 | TNFSF14 |
| ARHGAP9 | CSF3R | GCLC | KLK1 | NTRK3 | SEC63 | TNFSF15 |
| ARHGEF10 | CSH1 | GCLM | KLK12 | NUAK1 | SECISBP2 | TNFSF8 |
| ARHGEF11 | CSMD1 | GCM2 | KLK15 | NUDC | SEL1L | TNKS |
| ARHGEF12 | CSMD3 | GCNT1 | KLK3 | NUDT1 | SELE | TNNI2 |
| ARHGEF6 | CSNK1A1L | GCSH | KLK4 | NUDT6 | SELL | TNNT1 |
| ARHGEF7 | CSNK1D | GDAP1 | KLK7 | NUMA1 | SELP | TNNT3 |
| ARHGEF9 | CSNK2A2 | GDF15 | KLKB1 | NUMBL | SELPLG | TNP1 |
| ARID4A | CSNK2A3 | GDF9 | KMT5A | NUP155 | SEM1 | TNR |
| ARID4B | CSRP3 | GDI1 | KMT5B | NUP214 | SEMA4C | TNS2 |
| ARL11 | CSTB | GEMIN2 | KNG1 | NXF3 | SEMA4G | TNS3 |
| ARL14EP | CTF1 | GFAP | KPNA1 | NXF5 | SEMA6D | TNXA |
| ARL6IP5 | CTGF | GFI1 | KRT13 | NXNL1 | SEMA7A | TOMM40 |
| ARMS2 | CTH | GFI1B | KRT16 | OAS1 | SEMG1 | TOMM40L |
| ARPC3 | CTHRC1 | GFPT2 | KRT17 | OAS2 | SEPT12 | TOP1 |
| ARSE | CTNNA2 | GFRA1 | KRT18 | OAZ1 | SEPT9 | TOP1MT |
| ARSF | CTNNA3 | GFRA2 | KRT2 | OBSCN | SERPINA1 | TOP2A |
| ART4 | CTNND2 | GGH | KRT31 | OBSL1 | SERPINA10 | TOPBP1 |
| ARVCF | CTRC | GGT5 | KRT37 | OGG1 | SERPINA3 | TOX3 |
| AS3MT | CTSB | GH1 | KRT38 | OLFM2 | SERPINA6 | TP53AIP1 |
| ASAH2 | CTSC | GH2 | KRT4 | OLIG2 | SERPINA7 | TP53BP1 |
| ASCC1 | CTSG | GHRH | KRT6A | OLR1 | SERPINB11 | TP53I3 |
| ASCC3 | CTSZ | GHRHR | KRT6B | OPCML | SERPINB3 | TP73 |
| ASIP | CTTNBP2 | GHRL | KRT6C | OPLAH | SERPINB5 | TPCN2 |
| ASL | CUBN | GHSR | KRT75 | OPN4 | SERPINB6 | TPH1 |
| ASPRV1 | CUL2 | GIF | KRT8 | OPRD1 | SERPIND1 | TPH2 |
| ASS1 | CUL3 | GIMAP8 | KRT85 | OPRK1 | SERPINE1 | TPK1 |
| ASTN2 | CUL4B | GIP | KRT9 | OPRL1 | SERPINF1 | TPMT |
| ATF1 | CUL5 | GIPC3 | KRTAP1-1 | OPRM1 | SERPINF2 | TPO |
| ATF3 | CUL7 | GIPR | KRTCAP3 | OPTC | SERPINI2 | TPP2 |

| | | | | | | |
|---|---|---|---|---|---|---|
| ATG16L1 | CX3CR1 | GIT1 | KYNU | OR10X1 | SERTAD1 | TPRN |
| ATG7 | CXCL12 | GJA4 | L3MBTL1 | OR13G1 | SESN2 | TPTE |
| ATL1 | CXCL16 | GJC3 | LAMA4 | OR1B1 | SETDB2 | TRADD |
| ATP10A | CXCL5 | GJD2 | LAMA5 | OR51G1 | SEZ6 | TRAF3 |
| ATP10D | CXCR1 | GLCCI1 | LAMC1 | OR52H1 | SEZ6L2 | TRAF6 |
| ATP13A4 | CXCR3 | GLDC | LAMTOR2 | OR52N4 | SFTPA1 | TRAK2 |
| ATP1B1 | CXCR4 | GLMN | LARGE2 | OR5AC2 | SFTPA2 | TRAPPC10 |
| ATP2A1 | CYB5R4 | GLO1 | LBP | OR5H6 | SFTPB | TREH |
| ATP2A2 | CYBRD1 | GLP1R | LCE3B | OR8K3 | SFTPC | TRERF1 |
| ATP2A3 | CYCS | GLRA1 | LCK | ORC4 | SFTPD | TRHR |
| ATP2B4 | CYFIP1 | GLS | LCN10 | ORC6 | SGCA | TRIB1 |
| ATP2C1 | CYLD | GLTSCR1 | LCT | OTOA | SGCB | TRIB2 |
| ATP5E | CYP11A1 | GLUD1 | LDB1 | OTOF | SGCD | TRIB3 |
| ATP5SL | CYP11B1 | GLUD2 | LDB3 | OTOG | SGCE | TRIL |
| ATP6AP2 | CYP11B2 | GLUL | LDHA | OTOGL | SGCG | TRIM17 |
| ATP6V0A1 | CYP17A1 | GLYCTK | LDHB | OTOR | SGK1 | TRIM21 |
| ATP6V0A4 | CYP19A1 | GMIP | LDLRAD2 | OVCH2 | SGSH | TRIM22 |
| ATP6V1B1 | CYP1A1 | GMPS | LDLRAD4 | OVGP1 | SH2B1 | TRIM24 |
| ATP7A | CYP1A2 | GNA14 | LECT2 | OXCT1 | SH2D1A | TRIM33 |
| ATP8B1 | CYP21A2 | GNAI2 | LEF1 | OXTR | SH3GL1 | TRIM5 |
| ATPAF2 | CYP26A1 | GNAS-AS1 | LEFTY2 | P2RX1 | SHANK2 | TRIOBP |
| ATRIP | CYP26B1 | GNB1L | LEP | P2RX4 | SHBG | TRMU |
| ATRNL1 | CYP26C1 | GNS | LEPR | P2RX5 | SHMT1 | TROAP |
| ATXN3L | CYP27B1 | GOLGA3 | LFNG | P2RX7 | SHOX2 | TROVE2 |
| AURKA | CYP2A13 | GOLGA5 | LGALS13 | P2RY12 | SHROOM3 | TRPA1 |
| AURKC | CYP2A6 | GON4L | LGALS2 | P2RY4 | SI | TRPC3 |
| AVPR1A | CYP2B6 | GOPC | LGALS3 | PABPC4L | SIAE | TRPC4 |
| AVPR1B | CYP2C18 | GOSR2 | LGI1 | PACRG | SIGLEC12 | TRPC5 |
| AVPR2 | CYP2C19 | GOT1 | LGR5 | PADI4 | SIGLEC14 | TRPC6 |
| AXIN1 | CYP2C8 | GP2 | LHB | PAFAH1B3 | SIGLEC16 | TRPM2 |
| AXIN2 | CYP2C9 | GP6 | LHCGR | PAK3 | SIK2 | TRPM3 |
| AXL | CYP2D6 | GP9 | LHFPL5 | PALLD | SIM1 | TRPM4 |
| AZIN2 | CYP2E1 | GPAM | LHX1 | PAPD7 | SIM2 | TRPM6 |
| B2M | CYP2F1 | GPATCH8 | LHX8 | PAPSS2 | SIPA1 | TRPM7 |
| B3GALNT1 | CYP2G1P | GPBAR1 | LIAS | PARD3B | SIPA1L1 | TRPS1 |
| B3GALT1 | CYP2J2 | GPC6 | LIF | PARK2 | SIRT1 | TRPV1 |
| B3GALT2 | CYP2R1 | GPD1 | LIG1 | PARL | SIRT3 | TRPV5 |
| B3GALT5 | CYP2U1 | GPD1L | LIG3 | PARP1 | SIRT5 | TSG101 |

| | | | | | | |
|---|---|---|---|---|---|---|
| B3GAT1 | CYP2W1 | GPD2 | LIMK1 | PARP2 | SIX2 | TSHB |
| B3GAT2 | CYP3A4 | GPHN | LIN28A | PASK | SLA | TSHZ1 |
| B3GNT2 | CYP3A43 | GPI | LIN28B | PAWR | SLBP | TSLP |
| B3GNT3 | CYP3A5 | GPNMB | LIPA | PAX5 | SLC10A1 | TSPAN17 |
| B3GNT6 | CYP3A7 | GPR1 | LIPC | PAX8 | SLC10A2 | TSPAN7 |
| B3GNT7 | CYP46A1 | GPR12 | LIPE | PAX9 | SLC11A1 | TSPEAR |
| B4GALNT2 | CYP4A11 | GPR132 | LIPG | PC | SLC11A2 | TSPO |
| B4GALNT3 | CYP4A22 | GPR139 | LIPI | PCBD1 | SLC12A1 | TSSC4 |
| B4GALNT4 | CYP4B1 | GPR33 | LIPN | PCCA | SLC13A2 | TSSK4 |
| B4GALT1 | CYP4F12 | GPR55 | LITAF | PCCB | SLC14A1 | TTC14 |
| B4GALT2 | CYP4F2 | GPR68 | LLGL1 | PCDH11X | SLC14A2 | TTLL1 |
| B4GALT3 | CYP4F22 | GPS1 | LMAN1 | PCDH18 | SLC15A1 | TTLL11 |
| B4GALT4 | CYP4F3 | GPSM2 | LMBRD1 | PCDH19 | SLC16A1 | TUBA1A |
| B4GALT5 | CYP7A1 | GPX1 | LMF1 | PCDH9 | SLC16A3 | TUBA8 |
| B4GALT6 | CYS1 | GPX4 | LMNB2 | PCDHA1 | SLC17A1 | TUBB1 |
| BAALC | CYSLTR1 | GRB10 | LMO2 | PCDHA9 | SLC17A3 | TUBGCP5 |
| BAAT | CYSLTR2 | GREM1 | LMO4 | PCDHAC1 | SLC17A8 | TULP3 |
| BACE1 | D2HGDH | GRHL1 | LMTK3 | PCDHB4 | SLC18A1 | TXNIP |
| BAG3 | DAD1 | GRHPR | LNX1 | PCK2 | SLC19A1 | TXNRD2 |
| BANK1 | DAO | GRID1 | LNX2 | PCM1 | SLC1A1 | TYK2 |
| BARD1 | DAOA | GRIK1 | LOC10099684<br>2 | PCMT1 | SLC1A5 | TYMS |
| BARX2 | DAPK1 | GRIK3 | LOR | PCOLCE | SLC22A1 | TYMSOS |
| BAX | DAZL | GRIK4 | LOXHD1 | PCSK1 | SLC22A11 | TYRO3 |
| BAZ1B | DBI | GRIN3A | LOXL2 | PCSK2 | SLC22A12 | UACA |
| BCAM | DBT | GRK3 | LOXL3 | PCSK5 | SLC22A14 | UBA3 |
| BCAT1 | DCAF13 | GRK4 | LPA | PDCD5 | SLC22A18 | UBAC2 |
| BCAT2 | DCAF17 | GRK5 | LPAR1 | PDE10A | SLC22A18AS | UBE2B |
| BCHE | DCDC2 | GRM3 | LPIN1 | PDE11A | SLC22A2 | UBE2I |
| BCKDHA | DCK | GRM5 | LPIN2 | PDE12 | SLC22A3 | UBE2NL |
| BCKDHB | DCLK1 | GRM8 | LPIN3 | PDE4B | SLC22A4 | UBE3C |
| BCKDK | DCLRE1C | GRPR | LPP | PDE7B | SLC22A5 | UBN2 |
| BCL2 | DCP1B | GRXCR1 | LRCH1 | PDE8B | SLC22A6 | UBQLN2 |
| BCL2A1 | DCTD | GSDMA | LRFN5 | PDGFC | SLC22A9 | UBR3 |
| BCL2L1 | DCXR | GSDMB | LRP6 | PDGFRA | SLC23A1 | UBR7 |
| BCL2L11 | DDAH1 | GSE1 | LRP8 | PDGFRL | SLC24A2 | UCP1 |
| BCL2L2 | DDHD1 | GSK3B | LRRC6 | PDK1 | SLC25A12 | UCP2 |
| BCL6 | DDX20 | GSPT1 | LRRFIP2 | PDLIM3 | SLC25A3 | UCP3 |
| BCL9 | DDX25 | GSPT2 | LRSAM1 | PDLIM4 | SLC25A38 | UFD1L |

| | | | | | | |
|---|---|---|---|---|---|---|
| BCO1 | DDX3Y | GSTA1 | LRTOMT | PDLIM5 | SLC25A39 | UGCG |
| BDKRB2 | DDX5 | GSTA2 | LTBP1 | PDPK1 | SLC26A1 | UGGT1 |
| BDNF | DDX53 | GSTA3 | LTBP3 | PEAR1 | SLC26A3 | UGGT2 |
| BEX4 | DEC1 | GSTK1 | LTBR | PECAM1 | SLC26A4 | UGT1A10 |
| BHLHA9 | DECR1 | GSTM1 | LTF | PECR | SLC26A5 | UGT1A6 |
| BHLHE41 | DEF6 | GSTM3 | LTK | PEMT | SLC26A6 | UGT1A8 |
| BHMT | DEFB1 | GSTM4 | LTN1 | PENK | SLC26A9 | UGT2A1 |
| BICC1 | DEFB126 | GSTO1 | LUM | PER1 | SLC27A4 | UGT2A2 |
| BICD1 | DEFB4A | GSTO2 | LY96 | PER2 | SLC27A5 | UGT2B15 |
| BIRC5 | DENND4A | GSTP1 | LYN | PER3 | SLC28A1 | UGT2B17 |
| BLMH | DES | GSTT1 | LYZ | PFAS | SLC28A2 | UGT2B28 |
| BLVRA | DFNA5 | GSTT2B | LZTS1 | PFKM | SLC28A3 | UGT2B4 |
| BLZF1 | DFNB59 | GSTZ1 | MACROD2 | PGAM1 | SLC29A1 | UGT2B7 |
| BMP10 | DGAT1 | GTF2E1 | MAD1L1 | PGAM2 | SLC29A2 | UGT8 |
| BMP15 | DGCR14 | GTF2H1 | MAD2L1 | PGAM5 | SLC29A4 | UHRF1BP1 |
| BMP2K | DGCR5 | GTF2I | MAGEE2 | PGBD1 | SLC2A2 | UIMC1 |
| BMP5 | DGKD | GTF2IRD1 | MAGI2 | PGC | SLC2A4 | ULK4 |
| BMP7 | DGKE | GTF2IRD2 | MAGT1 | PGD | SLC2A9 | UMOD |
| BMPR1B | DHFR | GUCY2C | MAML2 | PGM1 | SLC30A10 | UNC13D |
| BMPR2 | DHH | GUCY2F | MAMLD1 | PGR | SLC30A2 | UNC5C |
| BOC | DHRS4L1 | GYG1 | MAN1A2 | PGRMC1 | SLC30A5 | UNC5CL |
| BPGM | DHTKD1 | GYG2 | MAN2A1 | PHB | SLC30A8 | UNC93A |
| BPI | DHX36 | GYPA | MAOA | PHEX | SLC31A1 | UNC93B1 |
| BPIFA1 | DIABLO | GYPB | MAOB | PHF11 | SLC34A1 | UNG |
| BRAP | DIAPH2 | GYPC | MAP2 | PHF2 | SLC34A2 | UNKL |
| BRCC3 | DIAPH3 | GYPE | MAP2K3 | PHF3 | SLC34A3 | UPB1 |
| BRD1 | DIO1 | GYS1 | MAP2K4 | PHF8 | SLC35D1 | UPF3B |
| BRD2 | DIO2 | GYS2 | MAP2K7 | PHKA1 | SLC35G2 | UPK3A |
| BRSK2 | DIP2A | GZMB | MAP3K1 | PHKA2 | SLC44A2 | UQCRB |
| BRWD1 | DIP2B | H2BFWT | MAP3K14 | PHKB | SLC46A1 | UROD |
| BRWD3 | DIP2C | H6PD | MAP3K15 | PHKG2 | SLC47A1 | USP1 |
| BSCL2 | DIRC2 | HABP2 | MAP3K8 | PHLPP2 | SLC47A2 | USP15 |
| BSG | DISC1 | HAL | MAP4K5 | PI3 | SLC4A1 | USP24 |
| BSND | DISP1 | HAMP | MAP6 | PICALM | SLC4A10 | USP26 |
| BST1 | DKK2 | HAND1 | MAP7D3 | PICK1 | SLC4A3 | USP46 |
| BTAF1 | DKK3 | HAND2 | MAPK10 | PIF1 | SLC4A7 | USP9Y |
| BTBD9 | DLC1 | HAPLN1 | MAPK8IP1 | PIGM | SLC52A1 | UST |
| BTC | DLEC1 | HARS2 | MARVELD2 | PIGR | SLC5A1 | UTF1 |

| | | | | | | |
|---|---|---|---|---|---|---|
| BTLA | DLG5 | HAS1 | MASP2 | PIGZ | SLC5A11 | UTP4 |
| BTN1A1 | DLGAP2 | HAVCR1 | MAST4 | PIK3C2G | SLC5A2 | UTRN |
| BTN2A1 | DLGAP3 | HAX1 | MASTL | PIK3C3 | SLC5A5 | UTS2 |
| BTRC | DLL3 | HBD | MAT1A | PIK3CB | SLC6A1 | UVSSA |
| C10orf105 | DLX3 | HBE1 | MATN3 | PIK3CG | SLC6A11 | VANGL1 |
| C10orf11 | DLX6 | HBEGF | MATR3 | PIK3R4 | SLC6A12 | VAPB |
| C12orf29 | DMBT1 | HBG2 | MAVS | PIM1 | SLC6A13 | VCAM1 |
| C12orf65 | DMC1 | HBM | MBD1 | PIN1 | SLC6A14 | VCL |
| C15orf62 | DMD | HBS1L | MBD3 | PIP4K2A | SLC6A18 | VCX3A |
| C16orf58 | DMGDH | HBZ | MBD4 | PIP5K1B | SLC6A2 | VDR |
| C1GALT1 | DMP1 | HCFC1 | MBL2 | PIP5K1C | SLC6A4 | VEGFA |
| C1GALT1C1 | DMRT1 | HCK | MBNL1 | PITPNA | SLC6A5 | VIP |
| C1QA | DMXL1 | HCLS1 | MC2R | PIWIL3 | SLC6A6 | VIPAS39 |
| C1QB | DNAJA4 | HCN2 | MC3R | PKD1 | SLC7A1 | VIPR2 |
| C1QC | DNAJB2 | HCN3 | MC4R | PKD1L1 | SLC7A10 | VKORC1 |
| C1S | DNAJB6 | HCN4 | MCC | PKD2 | SLC7A11 | VNN1 |
| C21orf91 | DNASE1L1 | HCRTR1 | MCCC1 | PKHD1 | SLC7A2 | VPREB1 |
| C2orf42 | DNASE2 | HCRTR2 | MCCC2 | PKLR | SLC7A5 | VPS33B |
| C3 | DNMT3L | HDAC9 | MCEE | PKM | SLC7A7 | VPS37A |
| C3AR1 | DOC2A | HDC | MCF2L2 | PKN3 | SLC7A9 | VPS54 |
| C4BPA | DOCK4 | HDLBP | MCFD2 | PKP1 | SLC8A1 | VRK1 |
| C5 | DOCK9 | HDX | MCHR1 | PLA2G2A | SLC9A3R1 | VSIG4 |
| C5AR2 | DOK1 | HELQ | MCL1 | PLA2G2D | SLC9A9 | VTN |
| C6 | DOK2 | HEPACAM | MCM4 | PLA2G4A | SLCO1A2 | VWF |
| C7 | DOK5 | HEPH | MCM6 | PLA2G4C | SLCO1B1 | WASF3 |
| C8A | DOLK | HES6 | MCM7 | PLA2G7 | SLCO1B3 | WDFY4 |
| C8B | DOLPP1 | HES7 | MCM9 | PLAG1 | SLCO1C1 | WDR13 |
| C9 | DOT1L | HEY1 | MCPH1 | PLAGL1 | SLCO2B1 | WDR3 |
| CA1 | DPCD | HEY2 | MDH1 | PLAT | SLCO5A1 | WDR62 |
| CA12 | DPP10 | HFE2 | MDM2 | PLAU | SLFN5 | WDR72 |
| CA6 | DPY19L2 | HGF | MDM4 | PLAUR | SLIT3 | WISP3 |
| CABIN1 | DPYS | HHEX | MDN1 | PLCB1 | SLITRK1 | WNK4 |
| CABP2 | DPYSL2 | HHIP | ME2 | PLCE1 | SLITRK5 | WNT4 |
| CACNA1C | DRD1 | HIF1A | MECOM | PLCZ1 | SLURP1 | WNT5B |
| CACNA2D1 | DRD2 | HIF1AN | MED17 | PLD2 | SMAD1 | WNT7A |
| CACNA2D3 | DRD3 | HINT1 | MEF2A | PLEKHG4 | SMAD2 | WWC1 |
| CACNB2 | DRD4 | HIP1 | MEGF10 | PLEKHG5 | SMAD5 | WWTR1 |
| CACNG3 | DRD5 | HIST3H3 | MEGF11 | PLIN1 | SMAD6 | XBP1 |

| | | | | | | |
|---|---|---|---|---|---|---|
| CACNG4 | DROSHA | HK2 | MEIS1 | PLIN4 | SMAD7 | XDH |
| CADM1 | DRP2 | HLX | MEP1B | PLOD2 | SMAD9 | XG |
| CADPS2 | DSC3 | HMBS | MEPE | PLSCR3 | SMAP1 | XIAP |
| CALCA | DSCAM | HMGA1 | MESDC2 | PLTP | SMARCA2 | XIST |
| CALCR | DSCR8 | HMGCL | MESP2 | PLXND1 | SMARCAD1 | XK |
| CALCRL | DSG1 | HMGCR | MEST | PMAIP1 | SMC1B | XKR4 |
| CALHM1 | DSG3 | HMGCS2 | MET | PML | SMG1 | XPNPEP2 |
| CALM1 | DSPP | HMMR | METTL21A | PMS2P3 | SMG6 | XRCC3 |
| CALM3 | DTNA | HMOX1 | MFAP4 | PNLIP | SMIM3 | XRCC5 |
| CALR3 | DUOX2 | HMOX2 | MFGE8 | PNP | SMN1 | XRCC6 |
| CALU | DUOXA1 | HMSD | MFSD2A | PNPLA2 | SMN2 | YARS |
| CAMK4 | DUSP23 | HMX2 | MGAT1 | PNPLA3 | SMNDC1 | YBX2 |
| CAMKK1 | DYM | HNF1B | MGAT3 | POF1B | SMOC2 | YTHDF2 |
| CAMKK2 | DYNAP | HNRNPH3 | MGAT4A | POFUT2 | SMPD3 | YWHAE |
| CAMKMT | DYSF | HOGA1 | MGAT4B | POLB | SMPX | ZAN |
| CAMP | DYX1C1 | HOMER2 | MGAT4C | POLE2 | SMUG1 | ZAP70 |
| CAMSAP2 | E2F1 | HOXA10 | MGAT5 | POLL | SMYD3 | ZBTB25 |
| CAPN10 | E2F4 | HOXA11 | MGAT5B | POLR2E | SNAPC4 | ZBTB40 |
| CAPN13 | E2F5 | HOXA2 | MGEA5 | POLR2F | SNAPC5 | ZBTB41 |
| CAPN3 | EBAG9 | HOXA3 | MGLL | POLR3H | SNCAIP | ZC3H3 |
| CARD11 | ECE1 | HOXA4 | MGMT | POLRMT | SNCB | ZC3HAV1 |
| CARD14 | ECE2 | HOXB6 | MGST2 | POMC | SND1 | ZCCHC12 |
| CARD8 | ECI1 | HOXC13 | MGST3 | POMP | SNORD116-10 | ZCCHC13 |
| CARD9 | ECM1 | HOXD10 | MIA3 | PON1 | SNORD50A | ZCCHC8 |
| CARTPT | ECSIT | HOXD13 | MIAT | PON2 | SNRK | ZDHHC15 |
| CASC16 | EDA | HOXD4 | MICAL1 | PON3 | SNTA1 | ZDHHC17 |
| CASP1 | EDA2R | HP | MIIP | POP1 | SNTG2 | ZDHHC24 |
| CASP12 | EDAR | HPD | MINPP1 | POSTN | SNX19 | ZDHHC6 |
| CASP2 | EDARADD | HPRT1 | MIPOL1 | POU4F3 | SNX3 | ZDHHC8 |
| CASP3 | EDN2 | HPSE2 | MIR146A | POU5F1 | SOCS1 | ZFAT |
| CASP5 | EEF1B2 | HR | MIR17HG | POU5F1B | SOCS3 | ZFHX3 |
| CASP8 | EEF2K | HRC | MIR206 | PPARA | SOD1 | ZFP36 |
| CASP9 | EEF2KMT | HRG | MIR510 | PPARD | SOD2 | ZFP36L1 |
| CAST | EFCAB5 | HRH2 | MIR96 | PPARG | SOD3 | ZFP36L2 |
| CAT | EFHC2 | HRH3 | MKL1 | PPARGC1A | SOGA3 | ZFP69 |
| CATSPER1 | EFNA5 | HS1BP3 | MLC1 | PPARGC1B | SOHLH1 | ZFP90 |
| CATSPER2 | EFR3A | HSD11B1 | MLLT10 | PPAT | SORBS1 | ZFYVE27 |
| CATSPER3 | EFTUD2 | HSD17B1 | MLLT3 | PPIA | SORCS1 | ZHX3 |

| | | | | | | |
|---|---|---|---|---|---|---|
| CATSPER4 | EGF | HSD17B2 | MLYCD | PPIG | SORL1 | ZIC4 |
| CAV3 | EGLN1 | HSD17B3 | MMAA | PPM1B | SORT1 | ZMYM3 |
| CBFB | EGR3 | HSD3B1 | MMAB | PPM1G | SOX17 | ZNF175 |
| CBLB | EHBP1 | HSD3B2 | MMEL1 | PPM1K | SOX18 | ZNF202 |
| CBR1 | EHD2 | HSF1 | MMP10 | PPOX | SOX7 | ZNF213 |
| CBR3 | EHHADH | HSP90AA1 | MMP12 | PPP1R1A | SP100 | ZNF224 |
| CBX2 | EIF3H | HSP90B1 | MMP13 | PPP1R3A | SP110 | ZNF24 |
| CBX4 | EIF4E | HSPA5 | MMP20 | PPP1R3C | SP7 | ZNF300 |
| CBY1 | EIF4H | HSPA8 | MMP3 | PPP2R1B | SP8 | ZNF350 |
| CCDC107 | ELAC2 | HSPA9 | MMP7 | PPP2R2C | SPAG16 | ZNF385B |
| CCDC12 | ELANE | HSPB1 | MMP8 | PPP3R1 | SPAG17 | ZNF41 |
| CCDC127 | ELAVL2 | HSPB3 | MMP9 | PRB1 | SPAG8 | ZNF419 |
| CCDC14 | ELF4 | HSPB7 | MNX1 | PRB3 | SPANXN5 | ZNF420 |
| CCDC170 | ELK1 | HSPB8 | MOCOS | PRB4 | SPATA13 | ZNF433 |
| CCDC50 | ELK3 | HTN3 | MOK | PRCC | SPATA16 | ZNF480 |
| CCDC66 | ELMOD2 | HTR1A | MPG | PRCP | SPATA21 | ZNF507 |
| CCDC78 | ELP2 | HTR1B | MPHOSPH8 | PRDM2 | SPATA31C1 | ZNF526 |
| CCDC8 | EME1 | HTR2A | MPI | PRDM9 | SPECC1 | ZNF592 |
| CCK | EMG1 | HTR2B | MPO | PRG4 | SPG20 | ZNF627 |
| CCKAR | EMX1 | HTR2C | MPP3 | PRH1 | SPG21 | ZNF674 |
| CCKBR | EMX2 | HTR3A | MPP4 | PRICKLE1 | SPI1 | ZNF711 |
| CCL11 | EN2 | HTR3B | MPP6 | PRICKLE2 | SPINK1 | ZNF75D |
| CCL17 | ENAM | HTR3C | MPP7 | PRKAA2 | SPINK5 | ZNF80 |
| CCL2 | ENO1 | HTR3E | MPST | PRKACA | SPP1 | ZNF804A |
| CCL22 | ENO3 | HTR5A | MR1 | PRKAG3 | SPRED2 | ZNF81 |
| CCL26 | ENSA | HTR6 | MRAP | PRKAR1B | SPRN | ZNHIT6 |
| CCL3 | ENTPD5 | HTR7 | MRC1 | PRKCA | SPRR3 | ZNRF1 |
| CCL5 | EOMES | HVCN1 | MREG | PRKCB | SPRY2 | ZPBP |
| CCL7 | EPB41 | IAPP | MRPL3 | PRKCH | SPTA1 | ZPBP2 |
| CCNA2 | EPB42 | IBSP | MRPL48 | PRKCSH | SPTAN1 | |
| CCNH | EPC2 | ICAM1 | MRPS22 | PRKD3 | SPTB | |
| CCPG1 | EPHA3 | ICAM4 | MRRF | PRKRA | SPTBN1 | |
| CCR1 | EPHA5 | ICAM5 | MS4A1 | PRL | SPTBN5 | |
| CCR2 | EPHA7 | ICK | MS4A12 | PRLH | SPTLC1 | |
| CCR3 | EPHB2 | ID3 | MS4A2 | PRLHR | SPTLC2 | |
| CCR5 | EPHB6 | ID4 | MS4A3 | PRLR | SRD5A2 | |
| CCR6 | EPHX1 | IDE | MS4A6A | PRM1 | SREBF2 | |
| CCR7 | EPO | IDH1 | MS4A6E | PRM2 | SREK1 | |

**Table S11.** List with 100 VUS pending on reclassification at the Genetics Department of the HU-FJD.

| ID | HGVSc | HGVSp | SYMBOL | AC | AF | AC_IRD | AF_IRD | AC_PC | AF_PC |
|---|---|---|---|---|---|---|---|---|---|
| chr1:94471103A>G | NM_000350.3: c.6041T>C | NP_000341.2: p.Met2014Thr | ABCA4 | 1 | 8.80E-05 | 1 | 2.90E-04 | 0 | 0.00E+00 |
| chr1:94502731A>C | NM_000350.3: c.3783T>G | NP_000341.2: p.Ser1261Arg | ABCA4 | 1 | 8.97E-05 | 1 | 2.90E-04 | 0 | 0.00E+00 |
| chr1:94506911G>C | NM_000350.3: c.3376C>G | NP_000341.2: p.Leu1126Val | ABCA4 | 1 | 8.77E-05 | 1 | 2.80E-04 | 0 | 0.00E+00 |
| chr1:94526199G>A | NM_000350.3: c.2054C>T | NP_000341.2: p.Thr685Ile | ABCA4 | 1 | 8.80E-05 | 1 | 2.80E-04 | 0 | 0.00E+00 |
| chr1:103352419G>T | NM_001190709.1: c.4685C>A | NP_001177638.1: p.Thr1562Asn | COL11A1 | 6 | 5.33E-04 | 3 | 8.60E-04 | 2 | 2.86E-04 |
| chr1:103474020T>C | NM_001190709.1: c.1565A>G | NP_001177638.1: p.Gln522Arg | COL11A1 | 1 | 8.98E-05 | 1 | 2.90E-04 | 0 | 0.00E+00 |
| chr1:150316692C>T | NM_001350529.1: c.1076C>T | NP_001337458.1: p.Thr359Met | PRPF3 | 2 | 1.81E-04 | 2 | 5.90E-04 | 0 | 0.00E+00 |
| chr1:186120331G>A | NM_031935.3: c.14609-1G>A | NA | HMCN1 | 1 | 8.97E-05 | 1 | 2.90E-04 | 0 | 0.00E+00 |
| chr1:197297911T>G | NM_001193640.2: c.430T>G | NP_001180569.1: p.Phe144Val | CRB1 | 14 | 1.26E-03 | 7 | 2.05E-03 | 5 | 7.26E-04 |
| chr1:197297973GGATGGAATT>G | NM_001193640.2: c.498_506del | NP_001180569.1: p.Ile167_Gly169del | CRB1 | 37 | 3.25E-03 | 22 | 6.24E-03 | 14 | 1.99E-03 |
| chr1:197298095T>C | NM_001193640.2: c.614T>C | NP_001180569.1: p.Ile205Thr | CRB1 | 38 | 3.33E-03 | 15 | 4.25E-03 | 23 | 3.26E-03 |
| chr1:202910771T>C | NM_001290553.1: c.1058A>G | NP_001277482.1: p.Tyr353Cys | ADIPOR1 | 1 | 9.03E-05 | 1 | 2.90E-04 | 0 | 0.00E+00 |
| chr1:215799146T>G | NA | NA | KCTD3 | 1 | 9.03E-05 | 1 | 2.90E-04 | 0 | 0.00E+00 |
| chr1:215848154C>T | NM_206933.3: c.13099G>A | NP_996816.2: p.Val4367Ile | USH2A | 1 | 8.77E-05 | 1 | 2.80E-04 | 0 | 0.00E+00 |
| chr1:215848921G>A | NM_206933.3: c.12332C>T | NP_996816.2: p.Ser4111Phe | USH2A | 1 | 8.77E-05 | 1 | 2.80E-04 | 0 | 0.00E+00 |
| chr1:215960035C>A | NM_206933.3: c.10364G>T | NP_996816.2: p.Ser3455Ile | USH2A | 1 | 8.77E-05 | 1 | 2.80E-04 | 0 | 0.00E+00 |
| chr1:216246603C>T | NM_206933.3: c.5612G>A | NP_996816.2: p.Gly1871Asp | USH2A | 9 | 7.90E-04 | 2 | 5.70E-04 | 6 | 8.50E-04 |
| chr1:216256830C>T | NM_206933.3: c.5266G>A | NP_996816.2: p.Val1756Ile | USH2A | 1 | 8.82E-05 | 1 | 2.90E-04 | 0 | 0.00E+00 |
| chr1:216258156G>A | NM_206933.3: c.5051C>T | NP_996816.2: p.Pro1684Leu | USH2A | 1 | 8.77E-05 | 1 | 2.80E-04 | 0 | 0.00E+00 |
| chr1:216500940T>G | NM_007123.5: c.841A>C | NP_009054.5: p.Thr281Pro | USH2A | 2 | 1.77E-04 | 1 | 2.90E-04 | 1 | 1.43E-04 |
| chr10:73498276G>A | NA | NA | C10orf105 | 3 | 2.63E-04 | 2 | 5.70E-04 | 1 | 1.42E-04 |
| chr10:85961599G>A | NM_001171971.3: c.562G>A | NP_001165442.1: p.Gly188Ser | CDHR1 | 1 | 8.88E-05 | 1 | 2.90E-04 | 0 | 0.00E+00 |
| chr10:85971932C>A | NM_001171971.3: c.1554-3C>A | NA | CDHR1 | 5 | 4.40E-04 | 2 | 5.70E-04 | 3 | 4.26E-04 |
| chr10:85971970C>G | NM_001171971.3: c.1589C>G | NP_001165442.1: p.Thr530Ser | CDHR1 | 4 | 3.57E-04 | 3 | 8.70E-04 | 1 | 1.44E-04 |
| chr10:102780410T>C | NM_001195263.2: c.893A>G | NP_001182192.1: p.Lys298Arg | PDZD7 | 1 | 8.93E-05 | 1 | 2.90E-04 | 0 | 0.00E+00 |
| chr11:61725731C>G | NM_001139443.2: c.648C>G | NP_001132915.1: p.Phe216Leu | BEST1 | 1 | 8.84E-05 | 1 | 2.90E-04 | 0 | 0.00E+00 |
| chr12:88448181G>A | NA | NA | C12orf29 | 1 | 8.98E-05 | 1 | 2.90E-04 | 0 | 0.00E+00 |

| | | | | | | | | | |
|---|---|---|---|---|---|---|---|---|---|
| chr12:88481670G>C | NM_025114.4: c.4081C>G | NP_079390.3: p.Leu1361Val | CEP290 | 1 | 8.85E-05 | 1 | 2.90E-04 | 0 | 0.00E+00 |
| chr12:88535042G>T | NA | NA | TMTC3 | 1 | 8.89E-05 | 1 | 2.90E-04 | 0 | 0.00E+00 |
| chr14:68196055C>G | NM_152443.3: c.806C>G | NP_689656.2: p.Ala269Gly | RDH12 | 1 | 8.79E-05 | 1 | 2.80E-04 | 0 | 0.00E+00 |
| chr14:88892973G>A | NM_001040428.3: c.674G>A | NP_001035518.1: p.Arg225His | SPATA7 | 3 | 2.69E-04 | 1 | 2.90E-04 | 2 | 2.88E-04 |
| chr16:57921818G>A | NM_001286130.2: c.3385C>T | NP_001273059.1: p.Arg1129Trp | CNGB1 | 1 | 8.90E-05 | 1 | 2.90E-04 | 0 | 0.00E+00 |
| chr16:57973358C>G | NM_001286130.2: c.1330G>C | NP_001273059.1: p.Glu444Gln | CNGB1 | 1 | 9.66E-05 | 1 | 3.30E-04 | 0 | 0.00E+00 |
| chr17:1554979T>G | NM_006445.4: c.6473A>C | NP_006436.3: p.His2158Pro | PRPF8 | 1 | 8.87E-05 | 1 | 2.90E-04 | 0 | 0.00E+00 |
| chr17:1558750T>C | NM_006445.4: c.5881A>G | NP_006436.3 :p.Ile1961Val | PRPF8 | 1 | 9.02E-05 | 1 | 2.90E-04 | 0 | 0.00E+00 |
| chr17:6328998C>A | NM_001033054.3: c.748G>T | NP_001028226.1: p.Ala250Ser | AIPL1 | 18 | 1.62E-03 | 6 | 1.75E-03 | 12 | 1.74E-03 |
| chr17:6337375G>C | NM_001033054.3: c.140C>G | NP_001028226.1: p.Thr47Arg | AIPL1 | 17 | 1.49E-03 | 6 | 1.70E-03 | 11 | 1.56E-03 |
| chr17:7906552G>A | NM_000180.4: c.187G>A | NP_000171.1: p.Ala63Thr | GUCY2D | 1 | 8.89E-05 | 1 | 2.90E-04 | 0 | 0.00E+00 |
| chr17:7917341G>C | NM_000180.4: c.2407G>C | NP_000171.1: p.Asp803His | GUCY2D | 1 | 9.00E-05 | 1 | 2.90E-04 | 0 | 0.00E+00 |
| chr17:7918305T>C | NM_000180.4: c.2705T>C | NP_000171.1: p.Val902Ala | GUCY2D | 1 | 8.77E-05 | 1 | 2.80E-04 | 0 | 0.00E+00 |
| chr17:26879514G>A | NM_001330166.2: c.-252C>T | NA | UNC119 | 1 | 9.04E-05 | 1 | 2.90E-04 | 0 | 0.00E+00 |
| chr17:58234860T>C | NM_000717.5: c.341T>C | NP_000708.1: p.Leu114Ser | CA4 | 10 | 9.01E-04 | 3 | 8.80E-04 | 6 | 8.70E-04 |
| chr17:79496011G>A | NM_001077182.3: c.454G>A | NP_001070650.1: p.Val152Met | FSCN2 | 1 | 8.77E-05 | 0 | 0.00E+00 | 1 | 1.42E-04 |
| chr17:79503252TGAA>T | NM_001077182.3: c.1071_1073del | NP_001070650.1: p.Lys357del | FSCN2 | 2 | 1.75E-04 | 0 | 0.00E+00 | 0 | 0.00E+00 |
| chr17:79503273C>T | NM_001077182.3: c.1085C>T | NP_001070650.1: p.Ala362Val | FSCN2 | 1 | 8.77E-05 | 1 | 2.80E-04 | 0 | 0.00E+00 |
| chr19:3770708C>T | NM_001319074.2: c.604G>A | NP_001306003.1: p.Ala202Thr | RAX2 | 1 | 8.93E-05 | 1 | 2.90E-04 | 0 | 0.00E+00 |
| chr19:7621417C>T | NM_001166111.2: c.3202C>T | NP_001159583.1: p.Arg1068Cys | PNPLA6 | 2 | 1.75E-04 | 1 | 2.80E-04 | 1 | 1.42E-04 |
| chr19:54628040G>A | NM_015629.4: c.855+5G>A | NA | PRPF31 | 1 | 8.82E-05 | 1 | 2.80E-04 | 0 | 0.00E+00 |
| chr2:29296025TGCTTGCCCA>T | NM_001029883.3: c.1094_1102del | NP_001025054.1: p.Leu365_Lys367del | C2orf71 | 2 | 1.75E-04 | 2 | 5.70E-04 | 0 | 0.00E+00 |
| chr2:98986517C>T | NM_001079878.2: c.79C>T | NP_001073347.1: p.Arg27Cys | CNGA3 | 1 | 8.88E-05 | 1 | 2.90E-04 | 0 | 0.00E+00 |
| chr2:99012444C>G | NM_001079878.2: c.757C>G | NP_001073347.1: p.Pro253Ala | CNGA3 | 15 | 1.32E-03 | 5 | 1.42E-03 | 7 | 9.93E-04 |
| chr2:99012834T>C | NM_001079878.2: c.1147T>C | NP_001073347.1: p.Ser383Pro | CNGA3 | 2 | 1.75E-04 | 2 | 5.70E-04 | 0 | 0.00E+00 |
| chr2:99013422G>A | NM_001079878.2: c.1735G>A | NP_001073347.1: p.Ala579Thr | CNGA3 | 2 | 1.75E-04 | 1 | 2.80E-04 | 1 | 1.42E-04 |
| chr2:112740548T>A | NM_006343.3: c.1274T>A | NP_006334.2: p.Val425Glu | MERTK | 1 | 8.78E-05 | 1 | 2.80E-04 | 0 | 0.00E+00 |
| chr2:112779018G>A | NM_006343.3: c.2209G>A | NP_006334.2: p.Val737Ile | MERTK | 1 | 8.77E-05 | 1 | 2.80E-04 | 0 | 0.00E+00 |
| chr2:112779920A>G | NM_006343.3: c.2435A>G | NP_006334.2: p.Tyr812Cys | MERTK | 3 | 2.63E-04 | 2 | 5.70E-04 | 1 | 1.42E-04 |

| | | | | | | | | | |
|---|---|---|---|---|---|---|---|---|---|
| chr2:182409512C>T | NM_001030311.2: c.1358G>A | NP_001025482.1: p.Gly453Glu | CERKL | 1 | 8.87E-05 | 0 | 0.00E+00 | 1 | 1.43E-04 |
| chr2:234217866G>A | NM_000541.5: c.31G>A | NP_000532.2: p.Glu11Lys | SAG | 20 | 1.80E-03 | 4 | 1.17E-03 | 14 | 2.03E-03 |
| chr20:3891453A>T | NM_001324191.2: c.338A>T | NP_001311120.1: p.Asn113Ile | PANK2 | 4 | 3.60E-04 | 3 | 8.70E-04 | 1 | 1.45E-04 |
| chr3:50231006C>A | NM_000172.4: c.359C>A | NP_000163.2: p.Ser120Ter | GNAT1 | 7 | 6.14E-04 | 2 | 5.70E-04 | 5 | 7.08E-04 |
| chr3:100962441ACAT>A | NM_016247.4: c.2731_2733del | NP_057331.2: p.Met911del | IMPG2 | 1 | 9.32E-05 | 1 | 3.00E-04 | 0 | 0.00E+00 |
| chr3:100964729T>A | NM_016247.4: c.1460A>T | NP_057331.2: p.His487Leu | IMPG2 | 2 | 1.75E-04 | 2 | 5.70E-04 | 0 | 0.00E+00 |
| chr3:100964889G>A | NM_016247.4: c.1300C>T | NP_057331.2: p.Pro434Ser | IMPG2 | 11 | 9.74E-04 | 5 | 1.43E-03 | 5 | 7.16E-04 |
| chr3:193366624A>T | NM_001354663.2: c.1442A>T | NP_001341592.1: p.Glu481Val | OPA1 | 1 | 9.03E-05 | 1 | 2.90E-04 | 0 | 0.00E+00 |
| chr4:619687G>T | NM_000283.3: c.272G>T | NP_000274.2: p.Arg91Leu | PDE6B | 1 | 8.89E-05 | 0 | 0.00E+00 | 0 | 0.00E+00 |
| chr4:6292935G>A | NM_001145853.1: c.472G>A | NP_001139325.1: p.Glu158Lys | WFS1 | 1 | 8.82E-05 | 1 | 2.80E-04 | 0 | 0.00E+00 |
| chr4:6303119C>T | NM_001145853.1: c.1597C>T | NP_001139325.1: p.Pro533Ser | WFS1 | 6 | 5.26E-04 | 4 | 1.13E-03 | 0 | 0.00E+00 |
| chr4:16026888C>T | NM_001145847.2: c.530G>A | NP_001139319.1: p.Arg177Gln | PROM1 | 1 | 8.97E-05 | 1 | 2.90E-04 | 0 | 0.00E+00 |
| chr6:42141500C>T | NM_000409.4: c.149C>T | NP_000400.2: p.Pro50Leu | GUCA1A | 37 | 3.30E-03 | 10 | 2.90E-03 | 24 | 3.45E-03 |
| chr6:42147099A>AGACGAGGAGGGGGCT | NM_000409.4: c.572_586dup | NP_000400.2: p.Glu191_Glu195dup | GUCA1A | 1 | 9.03E-05 | 1 | 2.90E-04 | 0 | 0.00E+00 |
| chr6:42672150G>A | NM_000322.5: c.781C>T | NP_000313.2: p.Leu261Phe | PRPH2 | 1 | 8.77E-05 | 1 | 2.80E-04 | 0 | 0.00E+00 |
| chr6:42672265GCAGGGC>G | NM_000322.5: c.660_665del | NP_000313.2: p.Pro221_Cys222del | PRPH2 | 2 | 1.75E-04 | 2 | 5.70E-04 | 0 | 0.00E+00 |
| chr6:42689961C>T | NM_000322.5: c.112G>A | NP_000313.2: p.Gly38Arg | PRPH2 | 1 | 8.77E-05 | 1 | 2.80E-04 | 0 | 0.00E+00 |
| chr6:65303156G>A | NM_001142800.2: c.3731C>T | NP_001136272.1: p.Thr1244Ile | EYS | 2 | 1.75E-04 | 2 | 5.70E-04 | 0 | 0.00E+00 |
| chr6:66205150C>G | NM_001142800.2: c.154G>C | NP_001136272.1: p.Asp52His | EYS | 1 | 8.93E-05 | 1 | 2.90E-04 | 0 | 0.00E+00 |
| chr6:66205279G>T | NM_001142800.2: c.25C>A | NP_001136272.1: p.Leu9Met | EYS | 1 | 8.93E-05 | 1 | 2.90E-04 | 0 | 0.00E+00 |
| chr6:70990715C>T | NM_001851.5: c.904G>A | NP_001842.3: p.Gly302Ser | COL9A1 | 6 | 5.41E-04 | 2 | 5.80E-04 | 4 | 5.81E-04 |
| chr7:23145649G>A | NM_001031710.3: c.4G>A | NP_001026880.2: p.Ala2Thr | KLHL7 | 1 | 8.77E-05 | 1 | 2.80E-04 | 0 | 0.00E+00 |
| chr7:128035018C>T | NM_000883.4: c.1475G>A | NP_000874.2: p.Arg492Gln | IMPDH1 | 6 | 5.41E-04 | 6 | 1.75E-03 | 0 | 0.00E+00 |
| chr7:128041130G>A | NM_000883.4: c.443C>T | NP_000874.2: p.Thr148Met | IMPDH1 | 2 | 1.78E-04 | 1 | 2.90E-04 | 1 | 1.43E-04 |
| chr8:10466978G>A | NM_178857.6: c.4630C>T | NP_849188.4: p.Arg1544Cys | RP1L1 | 1 | 1.05E-04 | 1 | 3.50E-04 | 0 | 0.00E+00 |
| chr8:10469047G>A | NM_178857.6: c.2561C>T | NP_849188.4: p.Pro854Leu | RP1L1 | 1 | 1.05E-04 | 1 | 3.50E-04 | 0 | 0.00E+00 |
| chr8:10480383G>C | NM_178857.6: c.329C>G | NP_849188.4: p.Pro110Arg | RP1L1 | 1 | 8.78E-05 | 0 | 0.00E+00 | 1 | 1.42E-04 |
| chr8:10480420C>T | NM_178857.6: c.292G>A | NP_849188.4: p.Asp98Asn | RP1L1 | 6 | 5.33E-04 | 3 | 8.60E-04 | 3 | 4.30E-04 |

| | | | | | | | | | |
|---|---|---|---|---|---|---|---|---|---|
| chr8:43025820C>T | NM_001363227.2: c.726C>T | NP_001350156.1: p.Ser242%3D | HGSNAT | 2 | 1.77E-04 | 1 | 2.90E-04 | 1 | 1.43E-04 |
| chr8:43046725C>T | NM_001363227.2: c.1237C>T | NP_001350156.1: p.Pro413Ser | HGSNAT | 7 | 6.14E-04 | 1 | 2.80E-04 | 6 | 8.50E-04 |
| chr8:55533586A>C | NM_006269.2: c.60A>C | NP_006260.1: p.Gln20His | RP1 | 1 | 8.80E-05 | 1 | 2.80E-04 | 0 | 0.00E+00 |
| chr8:55534144G>A | NM_006269.2: c.615+3G>A | NA | RP1 | 13 | 1.14E-03 | 5 | 1.42E-03 | 8 | 1.14E-03 |
| chr8:55538939T>C | NM_006269.2: c.2497T>C | NP_006260.1: p.Phe833Leu | RP1 | 2 | 1.80E-04 | 1 | 2.90E-04 | 0 | 0.00E+00 |
| chr8:87638255T>C | NM_019098.4: c.1534A>G | NP_061971.3: p.Ile512Val | CNGB3 | 12 | 1.06E-03 | 2 | 5.70E-04 | 9 | 1.28E-03 |
| chr8:97172796C>A | NM_001001557.4: c.125G>T | NP_001001557.1: p.Gly42Val | GDF6 | 3 | 2.63E-04 | 2 | 5.70E-04 | 1 | 1.42E-04 |
| chr9:2718887G>T | NM_133497.4: c.1148G>T | NP_598004.1: p.Arg383Leu | KCNV2 | 3 | 2.63E-04 | 1 | 2.80E-04 | 2 | 2.83E-04 |
| chrX:13753441T>G | NA | NA | TRAPPC2 | 2 | 1.77E-04 | 2 | 5.80E-04 | 0 | 0.00E+00 |
| chrX:18674836C>T | NM_000330.4: c.121G>A | NP_000321.1: p.Asp41Asn | RS1 | 2 | 1.75E-04 | 2 | 5.70E-04 | 0 | 0.00E+00 |
| chrX:38178172T>C | NM_000328.3: c.379A>G | NP_000319.1: p.Arg127Gly | RPGR | 1 | 9.02E-05 | 1 | 2.90E-04 | 0 | 0.00E+00 |
| chrX:41333211A>G | NM_022567.2: c.505A>G | NP_072089.1: p.Asn169Asp | NYX | 2 | 1.94E-04 | 2 | 6.30E-04 | 0 | 0.00E+00 |
| chrX:41333709C>T | NM_022567.2: c.1003C>T | NP_072089.1: p.Arg335Cys | NYX | 1 | 8.89E-05 | 1 | 2.90E-04 | 0 | 0.00E+00 |
| chrX:49062162G>A | NM_001256789.3: c.5584C>T | NP_001243718.1: p.Arg1862Cys | CACNA1F | 2 | 1.76E-04 | 1 | 2.80E-04 | 1 | 1.42E-04 |
| chrX:49067552T>A | NM_001256789.3: c.4261A>T | NP_001243718.1: p.Ile1421Phe | CACNA1F | 2 | 1.79E-04 | 2 | 5.80E-04 | 0 | 0.00E+00 |
| chrX:49068452G>C | NM_001256789.3: c.4009-3C>G | NA | CACNA1F | 2 | 1.76E-04 | 2 | 5.70E-04 | 0 | 0.00E+00 |

**Table S12.** Carrier frequency (in pseudocontrol cases) and frequency in cases with inherited retinal dystrophies for the top 10 genes with higher carrier frequency.

| Gene | Allele Count | Frequency | Frequency type |
|---|---|---|---|
| ABCA4 | 252 | 7,14 | Carrier |
| USH2A | 89 | 2,52 | Carrier |
| PDE6A | 36 | 1,02 | Carrier |
| CEP290 | 32 | 0,91 | Carrier |
| ADGRV1 | 27 | 0,76 | Carrier |
| CNGB3 | 22 | 0,62 | Carrier |
| EYS | 22 | 0,62 | Carrier |
| CRB1 | 20 | 0,57 | Carrier |
| RP1L1 | 20 | 0,57 | Carrier |
| NMNAT1 | 19 | 0,54 | Carrier |
| ABCA4 | 375 | 21,23 | IRD |
| USH2A | 264 | 14,95 | IRD |
| PDE6A | 36 | 2,04 | IRD |
| CEP290 | 33 | 1,87 | IRD |
| ADGRV1 | 36 | 2,04 | IRD |
| CNGB3 | 63 | 3,57 | IRD |
| EYS | 53 | 3,00 | IRD |
| CRB1 | 49 | 2,77 | IRD |
| RP1L1 | 30 | 1,70 | IRD |
| NMNAT1 | 12 | 0,68 | IRD |

### Supplementary Figures

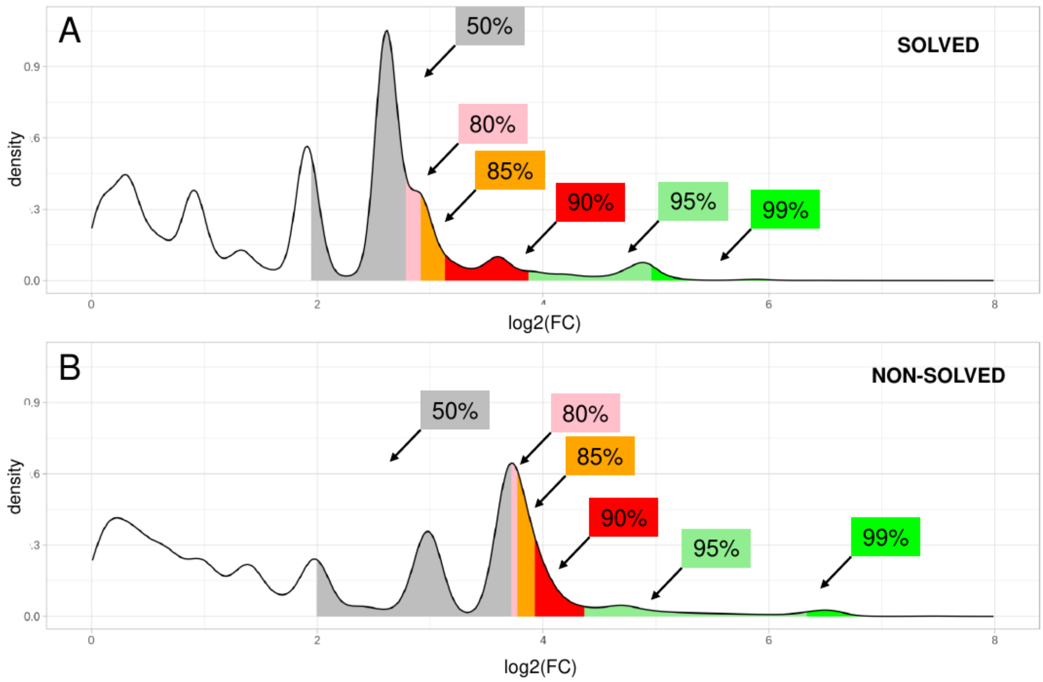


**Figure S1.** Distribution of the values of the fold changes (log2(FC)) calculated between the allelic frequencies in two IRD subcohorts: (A) solved and (B) non-solved, and the allelic frequencies in the pseudocontrols. Percentiles 50%, 80%, 85%, 90%, 95% and 99% are shown in both groups.

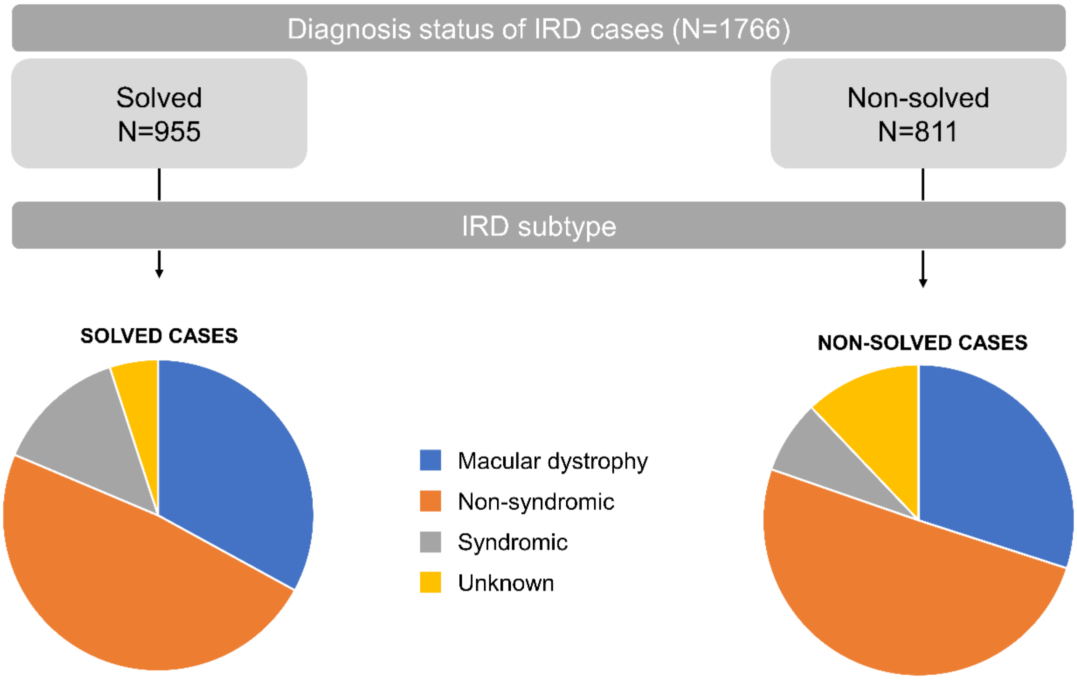


**Figure S2.** Description of the cohort of cases with inherited retinal dystrophies (IRD). Number of cases grouped by diagnostic status (solved and non-solved) and IRD subtype (syndromic, non-syndromic, macular dystrophies and unknown).

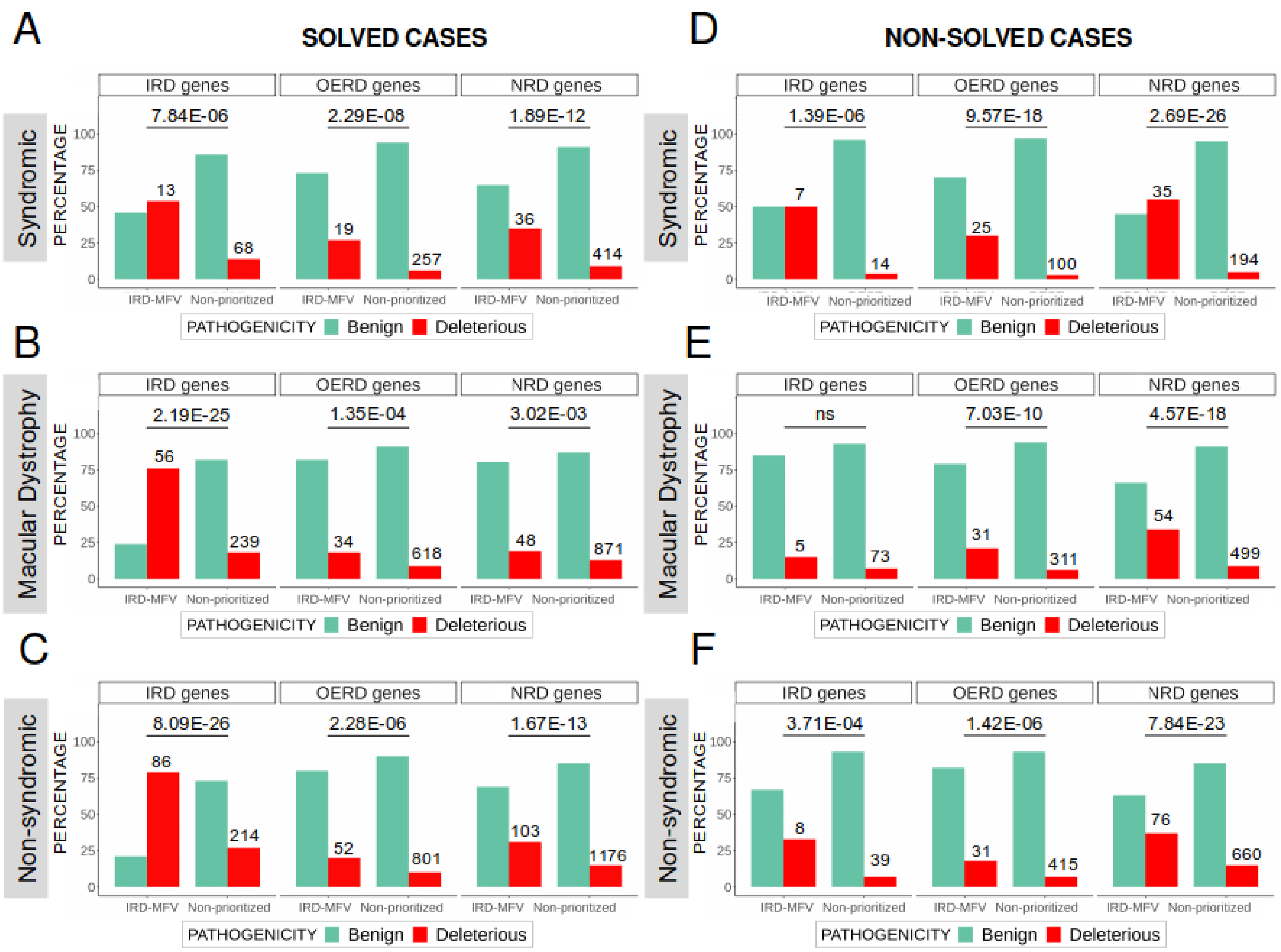


**Figure S3.** Proportion of deleterious and benign variants in both solved (A, B and C) and non-solved cases with inherited retinal dystrophies (D, E and F) for the different IRD subtypes: syndromic, non-syndromic and macular dystrophies. The p-values representing the enrichment of deleterious variants in IRD-MFVs are shown. The genes in which the IRD-MFVs are located are grouped in: inherited retinal dystrophies (RD genes), other eye related diseases (OERD genes) and other non-related diseases (NRD genes). Non-significant p-values are marked as “ns”.

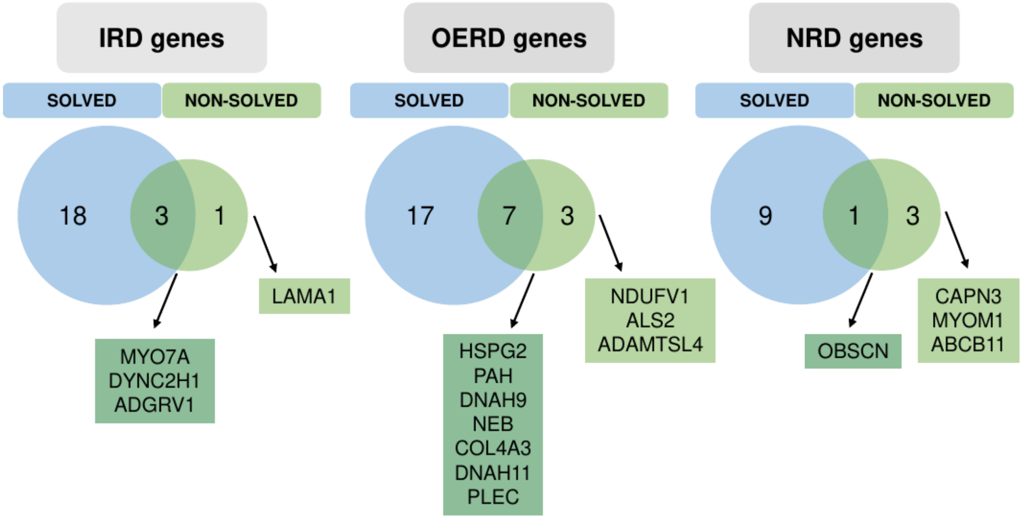


**Figure S4.** Intersection of genes prioritized in solved and non-solved cases with inherited retinal dystrophies. The genes are grouped as involved in: inherited retinal dystrophies (IRD genes), other eye related diseases (OERD genes) and other non-related diseases (NRD genes). We show only the names of the genes prioritized in non-solved IRD cases, in light green those unique to non-solved, and in dark green those in common with IRD solved cases.

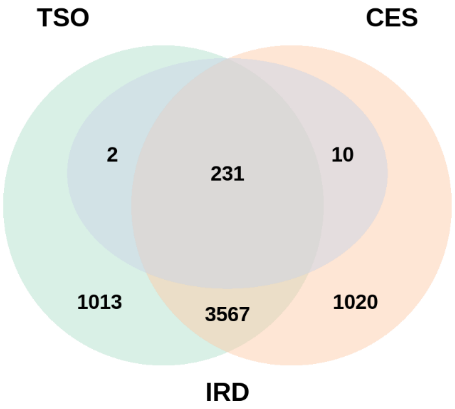


**Figure S5.** Intersection in genes included in the two clinical exomes used in the sequencing of the samples in the cohort: TruSightOne Sequencing Panel kit (TSO, Illumina, San Diego, CA), and Clinical Exome Solution Sequencing Panel kit (CES, Sophia Genetics, Boston, MA). Genes involved in inherited retinal dystrophies (IRD) are also highlighted.

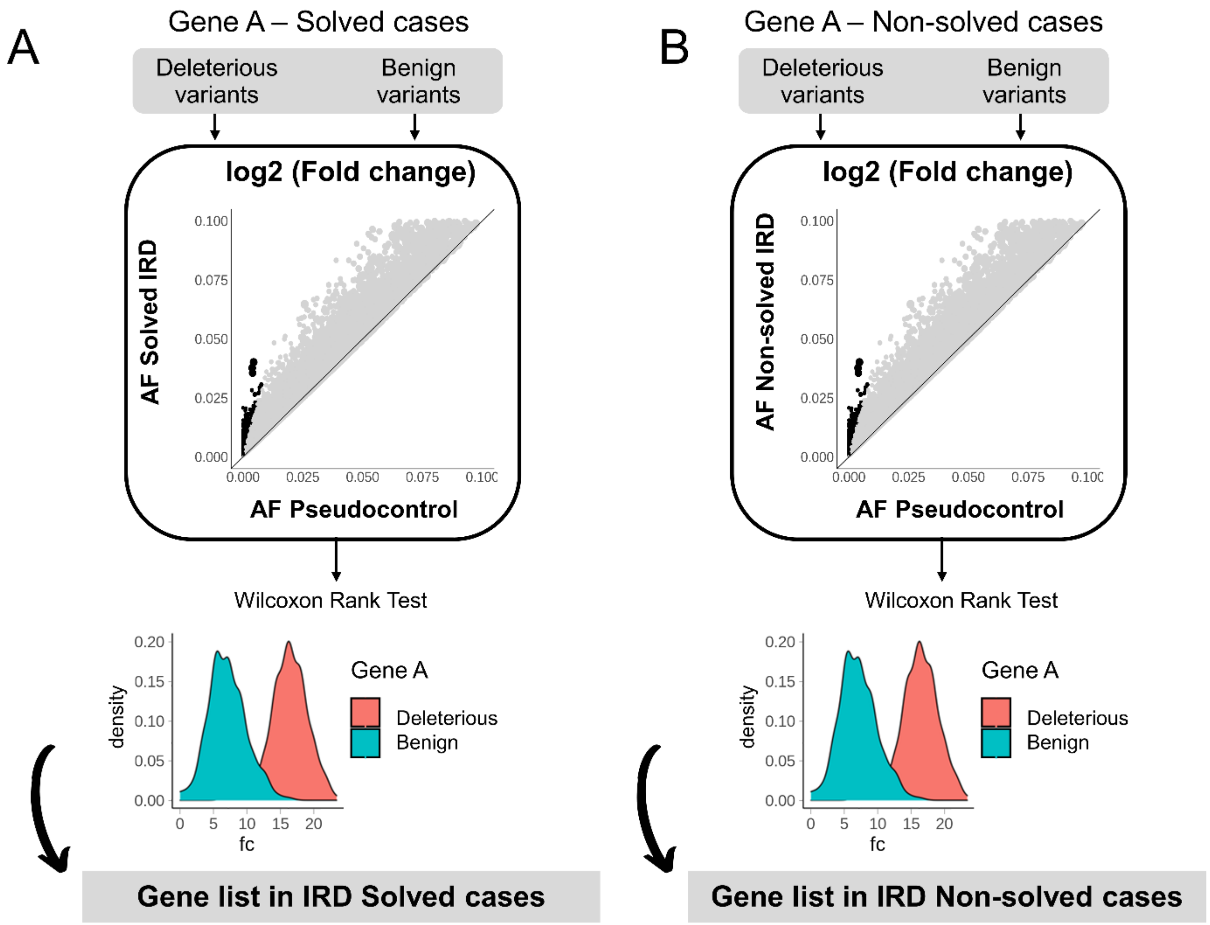


**Figure S6.** Workflow to perform gene prioritization in (A) solved and (B) non-solved cases with inherited retinal dystrophies (IRD).
